# Soluble TIM-3, likely produced by myeloid cells, predicts resistance to immune checkpoint inhibitors in metastatic clear cell renal cell carcinoma

**DOI:** 10.1101/2024.10.15.24315164

**Authors:** Ivan Pourmir, Nadine Benhamouda, Thi Tran, Hugo Roux, Joséphine Pineau, Alain Gey, Andyara Munoz, Nesrine Mabrouk, Nicolas Epaillard, Virginie Verkarre, Yann-Alexandre Vano, Eric Tartour, Stéphane Oudard

## Abstract

**Background and objectives:** Immunotherapies targeting PD-1 and CTLA-4 are key components of the treatment of metastatic clear cell renal cell carcinoma (mccRCC). However, they have distinct safety profiles and resistance to treatment can occur. We assess soluble TIM-3 (sTIM-3) in the plasma of mccRCC patients as a potential theranostic biomarker, as well as its source and biological significance.

**Methods:** We analyzed the association of sTIM-3 with overall survival (OS), tumor response, and common clinical and biological factors across two mccRCC cohorts treated with anti-PD-1 (n = 27), anti-PD-1 or anti-PD-1 + anti-CTLA-4 (n = 124). The origin and role of sTIM-3 are studied on tumor and blood samples, using multiplex immunohistochemistry and flow cytometry as well as a syngeneic tumor model with antitumor vaccination. We also reanalyzed publicly available single-cell transcriptomic (scRNAseq) data and mass cytometry data.

**Key findings and limitations:** sTIM-3 is elevated in the plasma of patients with mccRCC and shows distinct associations with survival on anti-PD-1 vs anti-PD-1 + anti-CTLA-4. sTIM-3 is independent from other clinical and biological factors. Myeloid immune cells appear as the prominent source of sTIM-3, which may indicate their dysfunctional role in the antitumor immune response. Future investigations are warranted in patients treated with anti-PD-1 + antiangiogenic therapies. Further functional studies are needed to confirm its theranostic value and clarify its role in the immune response.

**Conclusions and clinical implications:** sTIM-3 appears to be a promising biomarker for optimizing treatment strategies in ccRCC as well as a potential therapeutic target.

## Introduction

Immune checkpoint inhibitors (ICI) were introduced in clear cell renal cell carcinoma (ccRCC) as second line in the metastatic setting (mccRCC) with the approval of nivolumab monotherapy, an antibody blocking the-programmed cell death protein-1 (PD-1) ^1^. Anti-PD-1 were later combined in the first line with ipilimumab, an anti-cytotoxic T-lymphocyte-associated protein 4 (anti-CTLA-4) or antiangiogenic tyrosine kinase inhibitors (TKI) ^2^. The International Metastatic RCC Database Consortium (IDMC) score is the only validated theranostic biomarker for mccRCC and previously helped selecting nivolumab + ipilimumab (N+I) over sunitinib ^3,4^. No predictive biomarkers exist to discern patients who may achieve maximal efficacy with N+I rather than other anti-PD-1-based regimens.

T cell immunoglobulin and mucin domain-containing molecule 3 (TIM-3) is an immunomodulatory transmembrane protein classically ascribed as exhaustion marker on T cells, conferring them dysfunctionality ^5^. In ccRCC, TIM-3-expressing T cells have been associated with a poor prognosis ^6,7^ and resistance to anti-PD-1 as monotherapy ^8,9^ or combined with antiangiogenics ^10^. Clinical trials targeting TIM-3 in mccRCC are underway ^11,12^. Soluble isoforms of TIM-3 (sTIM-3) are engendered through proteolytic cleavage ^13,14^. Blood-based biomarkers possess the advantages of ready accessibility and proximity to baseline measurements. We assessed whether plasma sTIM-3 is associated with resistance of mccRCC to ICI, given the detrimental value of TIM-3 expression in the TME ^15^ and sTIM-3 association with prognosis regardless of treatment ^16^.

Here we demonstrate acute sTIM-3 increase induced via antitumor immunization in mice. We assess chronic plasma sTIM-3 elevation in mccRCC patients, its association with outcome under ICI. We investigate sources of sTIM-3 with analyses of tumor specimens and peripheral blood mononuclear cells (PBMC) from mccRCC patients.

## Material and methods

### Patient cohorts and sample collection

ICI-treated mccRCC: Colcheckpoint (nivolumab monotherapy after prior antiangiogenic) and BIONIKK (first line nivolumab monotherapy or N+I ^17^) independent cohorts approved by the French Health authorities and ethics committee [CPP Ile-de-France 8 (ref.16.10.69) and CPP Ouest I SI CNRIPH n18.11.21.67518 respectively]. Healthy adult human samples were drawn from blood donation. Plasma and PBMC were collected just before ICI initiation for mccRCC. Multiplex immunohistochemistry (IHC) was performed on formalin-fixed, paraffin-embedded (FFPE) primary tumors. Participants provided informed written consent.

### Mouse models

TC-1 cells were obtained from TC Wu’s laboratory (John Hopkins Hospital) and injected into anesthetized animals. Mice were vaccinated with 200µg G15F peptide from the HPV-16 E7 protein (Genosphere Biotechnologies) and 2µg of α-galactosyl-ceramide (α-Gal-Cer) adjuvant (Funakoshi Co.).

CD8+ T cells depletion was done *in vivo* by intraperitonel injections of a depleting antibody (100µg/100µl anti-mouse CD8α rat IgG2bκ, clone 2.43 InVivoMab™, BioXCell).

Experiments were conducted on 8-week-old female C57BL/6J mice (Janvier Labs) and approved by the ethics committee of the University Paris Cité (CEEA34).

### sTIM-3 quantification

sTIM-3 was quantified in human plasma samples using the Luminex® Multiplex ProcartaPlex^TM^ Human Immuno-Oncology Checkpoint Marker Panel 1 (14-Plex) kit (Thermofisher®), following the manufacturer’s instructions. Measures were acquired on a BioPlex-200 (BioRad®).

sTIM-3 was quantified in mouse serum by ELISA (Mouse Tim-3 SimpleStep ELISA® kit - ab255721 Abcam®), absorbance was measured on a Spectrostar microplate reader and converted to concentrations using the BMG Labtech software.

### Multiplex IHC

FFPE slides were incubated with specific antibodies (details in Supp.Table 2.A) and stained in a BOND- RX automate (Leica®) using Opal^TM^ secondary antibodies and fluorescent reagents. Specificity of TIM-3 was confirmed on additional ccRCC samples with the corresponding isotype control. Images were acquired in a Vectra Polaris scanner (Akoya/PerkinElmer) and analyzed with HALO^TM^ (v.3.6.4, IndicaLab®).

### Flow cytometry

PBMC were isolated by ficoll^TM^ density gradient centrifugation and frozen. PBMC were thawed, marked for viability (Zombie NIR, BioLegend®) and stained with antibodies against human markers (details in Supp.Table 2.B). Acquisition was performed on a Navios 10 colors cytometer (Beckman Coulter®), analyses were performed in Kaluza® (v 2.1). Thresholds for TIM-3 and CD14 positivity were set with corresponding control isotypes.

### Single-cell RNA sequencing (scRNAseq) data analyses

ScRNAseq data from ccRCC and non-cancerous human kidneys were obtained from publicly available datasets ^18–22^ and analyzed in R (v.4.1.1) using Seurat (v.5.0.3). Cells were annotated through SingleR (v.1.8.1) and manually. Differential gene expression (DEG) and Gene set enrichment (GSEA) analyses were performed using MAST (v.1.20.0) and enrichR (v.3.2). Pseudobulk analysis of scRNAseq data was performed with DESeq2 (v.1.34.0) on aggregated transcripts counts.

### Statistical analysis

Comparisons were performed for quantitative continuous variables using Mann-Whitney U (a.k.a. Wilcoxon ranks sum), Kruskal–Wallis, paired Wilcoxon, Friedman and Student’s t tests. Overall survival (OS) probabilities were estimated by the Kaplan-Meier method, differences were quantified by Cox hazard ratios (HR) and tested using the log-rank. Two-tailed p-values <0.05 were considered statistically significant. Analyzes were performed using the R software (v.4.3.2; R Foundation for Statistical Computing, Vienna, Austria).

## Results

### Plasma sTIM-3 is elevated in mccRCC

Plasma sTIM-3 levels were increased in treatment-naive mccRCC patients (median = 110.33 pg/L, IQR 65.36 to 151.10) compared to healthy adults (median = 46.10 pg/L, IQR 40.81 to 50.42, p < 10^-3^) (**Fig.1.A**).

**Fig. 1:**
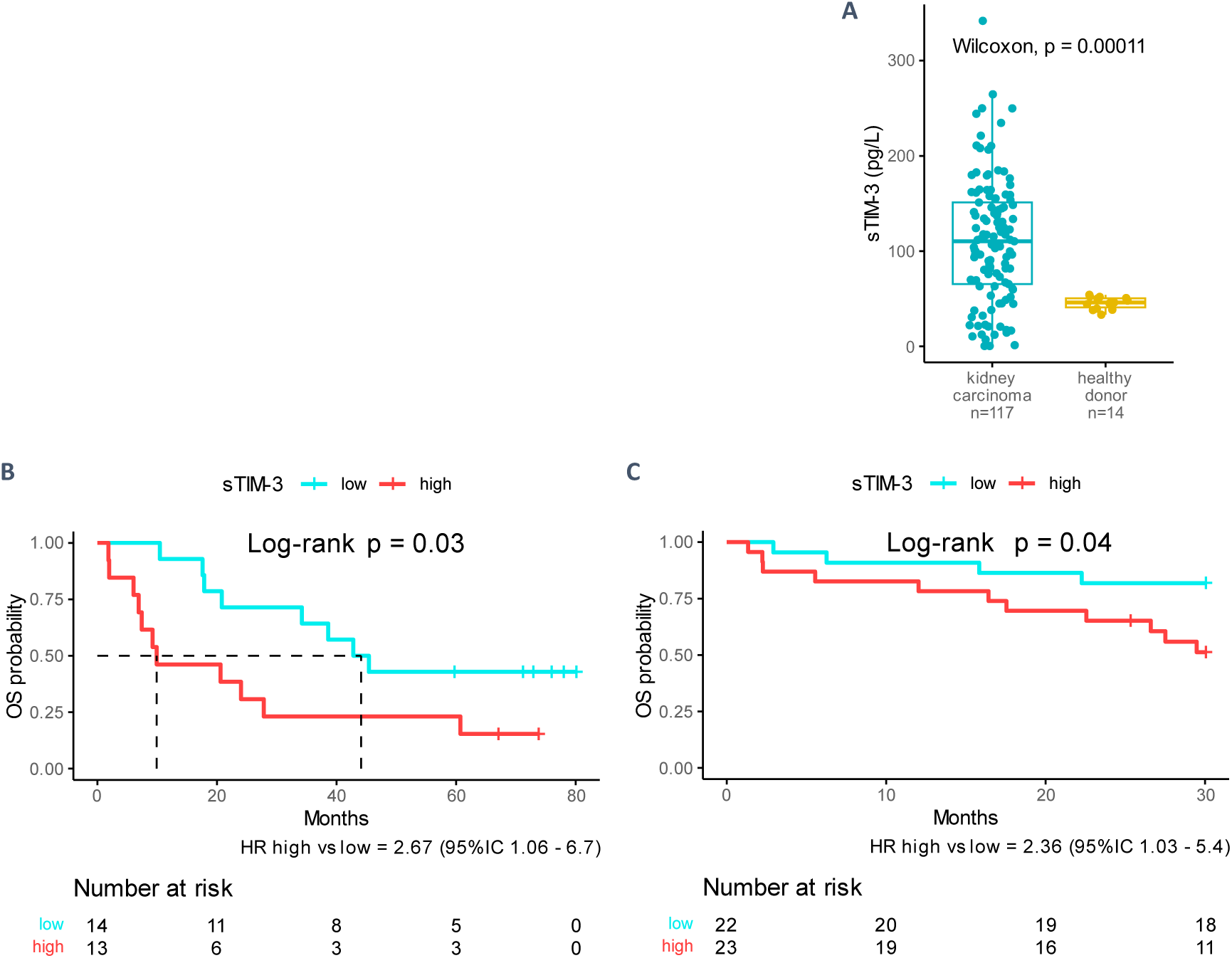
sTIM-3 plasma levels in ccRCC patients and association with OS_A_ under anti-PD1. A. sTIM-3 plasma levels were measured in treatment-naive ccRCC patients from the BIONIKK cohort before ICI treatment, and in healthy adults. B & C. Patients were categorized in sTIM-3 high or low, depending on whether their individual values of plasma sTIM-3 fell below or above the median value within each cohort. OS was estimated using the Kaplan-Meier method and differences between sTIM-3-low and -high groups were test with the log rank method in the Colcheckpoint cohort (A, n=27) and the BIONIKK cohort (B, n=45), patients’ characteristics are found in Supplementary Table 1.

To assess sTIM-3 at the early stage of the tumor response, we used a syngeneic TC-1 mouse tumor model, spontaneously poorly immunogenic but for which a specific anti-tumor immune response is elicited via vaccination. Control mice solely inoculated with TC-1 cells (day 0 – D0) showed no significant variation of plasma sTIM-3 18 days (D18) after the graft (D0 to D18 mean variation = -29.7 pg/L), whereas vaccinated mice exhibited a significant increase (D0 to D18 mean variation = +735.2 pg/L, p = 0.015 control versus vaccine) (**Supp. Fig.2**), suggesting a prominent role for the immune response in the production of sTIM-3.

### sTIM-3 is associated with OS in mccRCC treated with ICI

High baseline sTIM-3 (above the median within each cohort) was associated with poor OS on nivolumab monotherapy in both the Colcheckpoint (n = 27, HR for death sTIM-3-high vs sTIM-3-low = 2.67, p = 0.03) and BIONIKK (n = 45, HR sTIM-3-high vs sTIM-3-low = 2.36, p = 0.04) cohorts (**Fig.1.B & C**).

There was no difference according to sTIM-3 stratification in patients from the BIONIKK trial treated with N+I (**Supp.Fig.3.A**). Furthermore, we found that sTIM-3-low participants of the nivolumab monotherapy and N+I arms had comparable OS, whereas for sTIM-3-high participants, N+I was significantly superior to nivolumab (**Supp.Fig.3.B**). To verify that results in the N+I group were not confounded by IMDC scores (**Supp.Table 1.C**), a Cox model including IMDC and sTIM-3 categories was constructed. Still, the adjusted analysis evidenced no relationship between sTIM-3 category and OS under N+I (HR high vs low = 1.0, 95% IC 0.40 - 2.72) (**Supp.Fig.4**).

This suggests that sTIM-3 can distinguish mccRCC patients likely to achieve prolonged survival under anti-PD-1 monotherapy, while the anti-PD-1 + anti-CTLA4 combination retains efficacy in sTIM-3-high patients.

### sTIM-3 is independent from clinical and biological prognostic markers

IMDC categorization was not associated with sTIM-3 (**Fig.2.A**) and sTIM-3 was neither correlated with systemic inflammation markers included in the score (neutrophils and platelets counts, decreased hemoglobin) nor with other biomarkers (**Suppl.Fig.5** and **Supp.Fig.7)**.

**Fig. 2:**
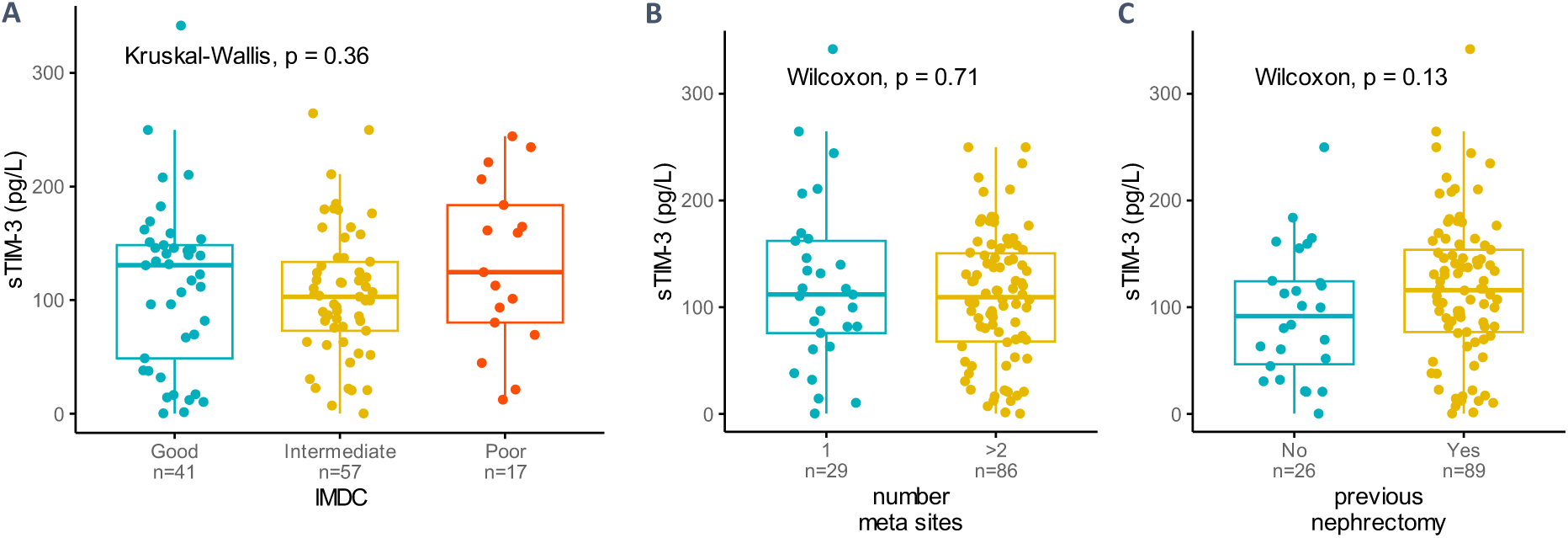
Comparison of sTIM-3 plasmatic levels in participants of BIONIKK (n= 133) categorized on IMDC score and tumor burden. A. sTIM-3 versus IMDC score calculated at baseline before ICI initiation. B. sTIM-3 versus number of metastatic sites at baseline (1 site or ≥2 sites). C. sTIM-3 versus history of primary tumor removal (previous nephrectomy).

Unlike CRP (**Supp.Fig.6**), sTIM-3 was not associated with tumor burden proxies (number of metastatic sites or presence of the primary tumor, **Fig.2.B**&**C**). This suggests another source than tumor cells for sTIM-3, consistent with the mouse model, and that sTim-3 does not simply reflect a higher tumor burden or systemic inflammation.

### Plasma sTIM-3 is likely produced by myeloid cells in ccRCC

#### A. *In situ* multiplex IHC of ccRCC tumors

We assessed the expression of TIM-3 by tumor cells - pancytokeratin and/or PAX8 positive - and tested its association with plasma sTIM-3 in a subset of BIONIKK participants (n = 22) (**Fig.3.A**).

**Fig. 3:**
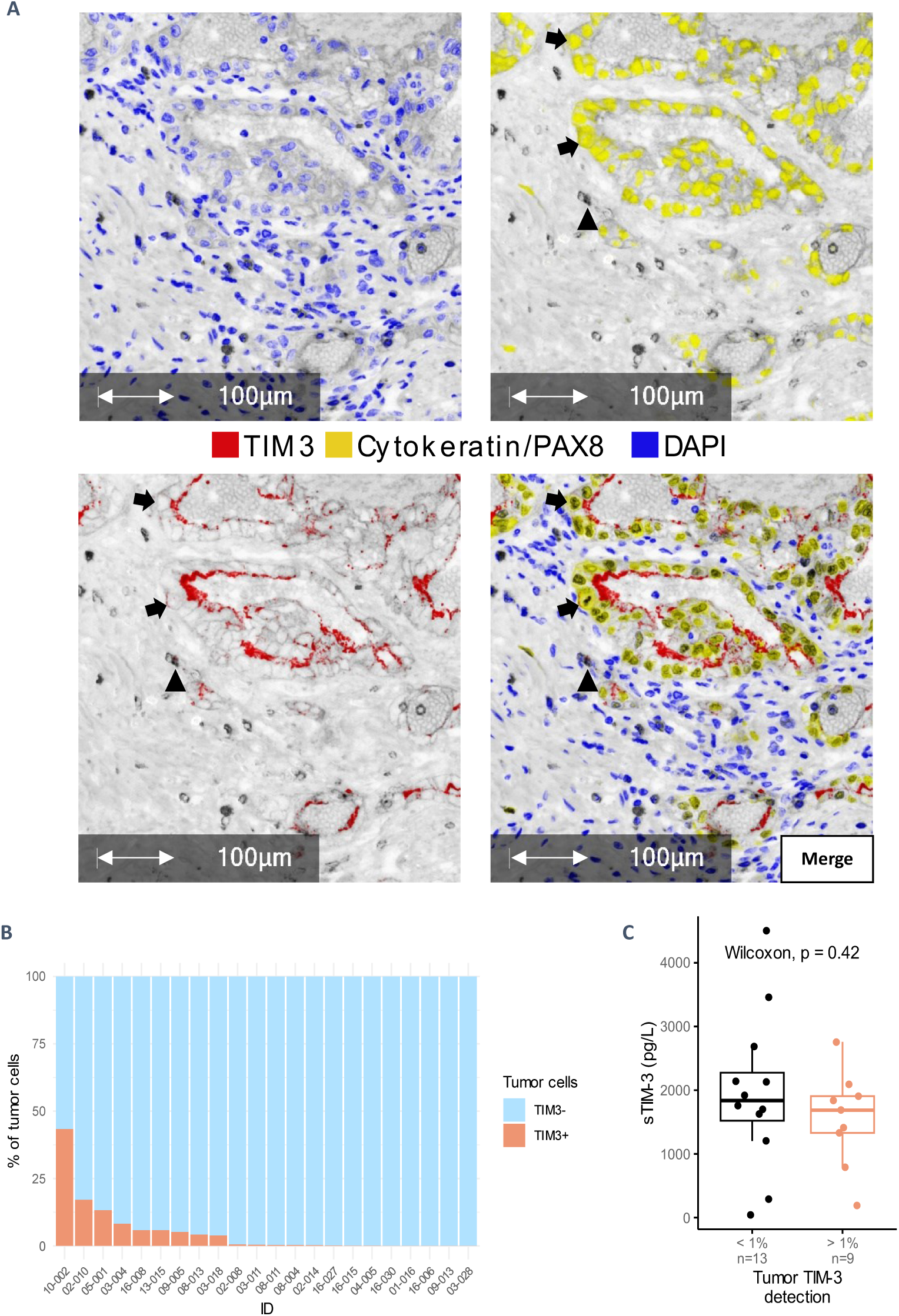
TIM-3 IHC detection on ccRCC tumors. 10 random tumor regions evenly spread on the slides were selected for analyzes. A custom phenotyping algorithm was used for quantification of total PAX8+Cytokeratin+ tumor cells and manual counting was performed for quantification of TIM3+ tumor cells. A. Absorption view (x20) of a ccRCC primary tumor FFPE sample analyzed with multiplex IHC. Blue: DAPI; Yellow: PanCytokeratine-PAX; Red: TIM-3. Arrows: TIM-3 positive tumor cells; Arrowhead: TIM-3 positive non-tumor cell. B. % of TIM-3-positive tumor cells among tumor cells in each sample (n = 22). C. Association of plasma sTIM-3 with TIM-3 tumor status.

There was a high intra-tumor and inter-samples variability of TIM-3 staining of tumor cells (**Fig.3.B**), with a median of 0.34% (IQR 0.05% to 5.61%) positive tumor cells. Tumors with >1% TIM-3 positive tumor cells were classified as TIM-3 positive. TIM-3 positivity was neither associated with higher levels of plasma sTIM-3 (**Fig.3.C**), nor with survival under nivolumab monotherapy (**Supp.Fig.8**).

TIM-3-positive non-tumor cells did not appear to be correlated to plasma sTIM-3 (Spearman correlation = -0.26, p = 0.25, **Supp.Fig.9**), leading us to investigate circulating immune cells.

#### B. scRNAseq of ccRCC tumors

The main hypothesis for the production of sTIM-3 in human is its shedding from double-positive HAVCR2+ADAM+ cells, co-expressing HAVCR2 (encoding TIM-3) and one or both of the metalloproteinases (ADAM10 and ADAM17) for which a cleaving activity of membrane TIM-3 is reported ^13,14^. We quantified the expression of these genes in several scRNAseq datasets, results are shown on Obradovic et al. dataset (**Fig.4**).

**Fig. 4.**
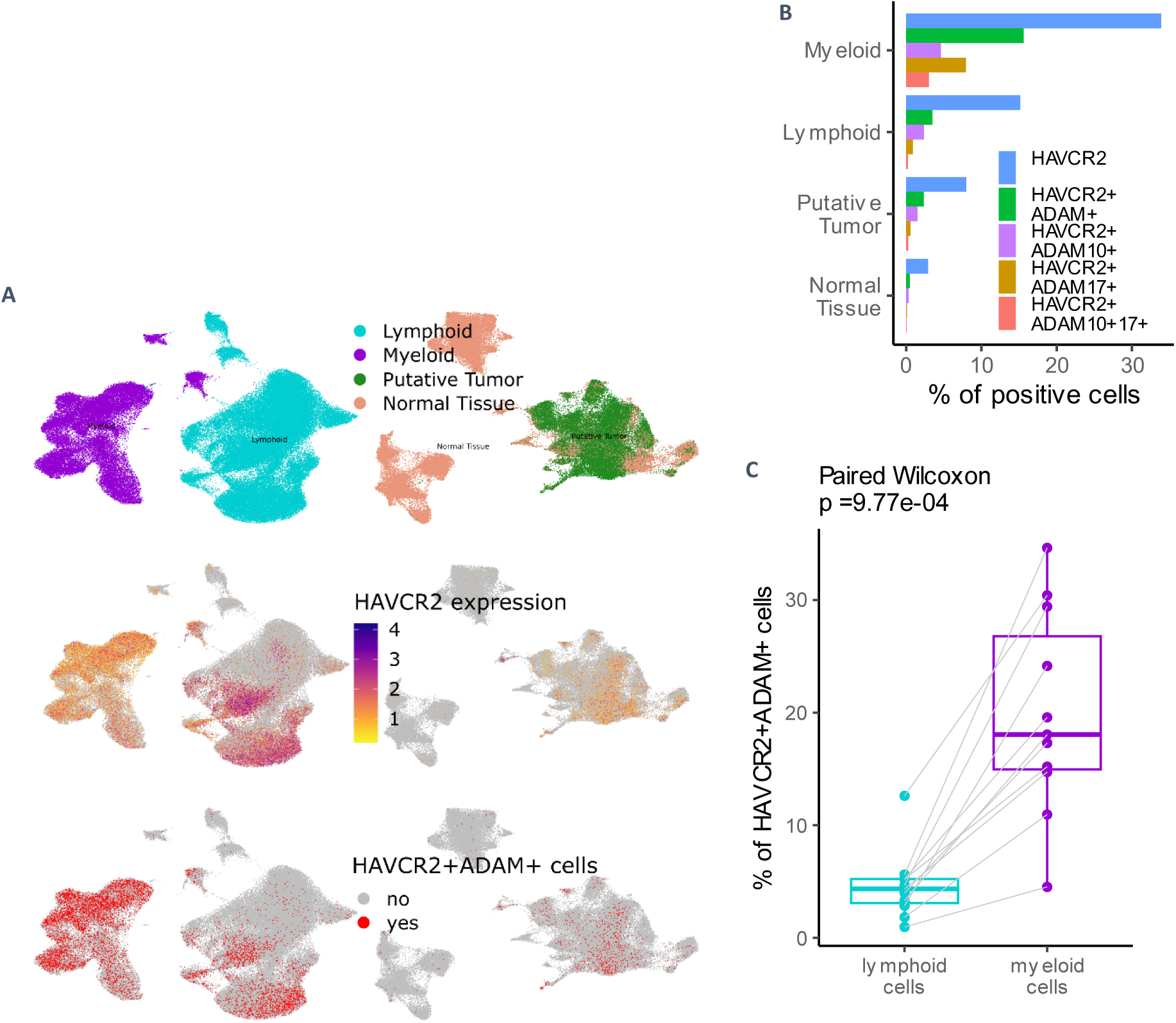
Obradovic et al. ccRCC scRNAseq dataset. A. Tumor and adjacent tissue **B** samples. Upper section: UMAP of cell lineages; middle section: HAVCR2 expression; lower section: HAVCR2+ADAM+ double-positive cells repartition. B. % of cells in the pooled dataset expressing HAVCR2 and ADAM10 and/or ADAM17 in broad lineages. HAVCR2+ADAM10+ - detection of ADAM10 transcripts but not ADAM17; HAVCR2+ADAM17+ - detection of ADAM17 transcripts but not ADAM10, HAVCR2+ADAM10+17+ - detection of both metalloproteinases. C.Comparison of the proportions of HAVCR2+ADAM+ cells within the lymphoid and myeloid lineages, for each patient in tumor samples.

Although some CD8+ and NK lymphocytes clusters had high transcription levels of HAVCR2, myeloid clusters (**Supp.Fig.10<u>,11</u> & <u>12</u>**) were identified as the most enriched lineage in HAVCR2+ADAM+ cells, with even distributions of the 3 different double-positive types (**Fig.7.A** & **B**; **<u>Supp.Fig.10 & 12</u>**). HAVCR2 expression was detected in renal tissue clusters but they contained marginal proportions of HAVCR2+ADAM+ cells (**Fig.5.A** & **B**). Consistent findings were reproduced on 3 other ccRCC scRNAseq datasets.

**Fig. 5:**
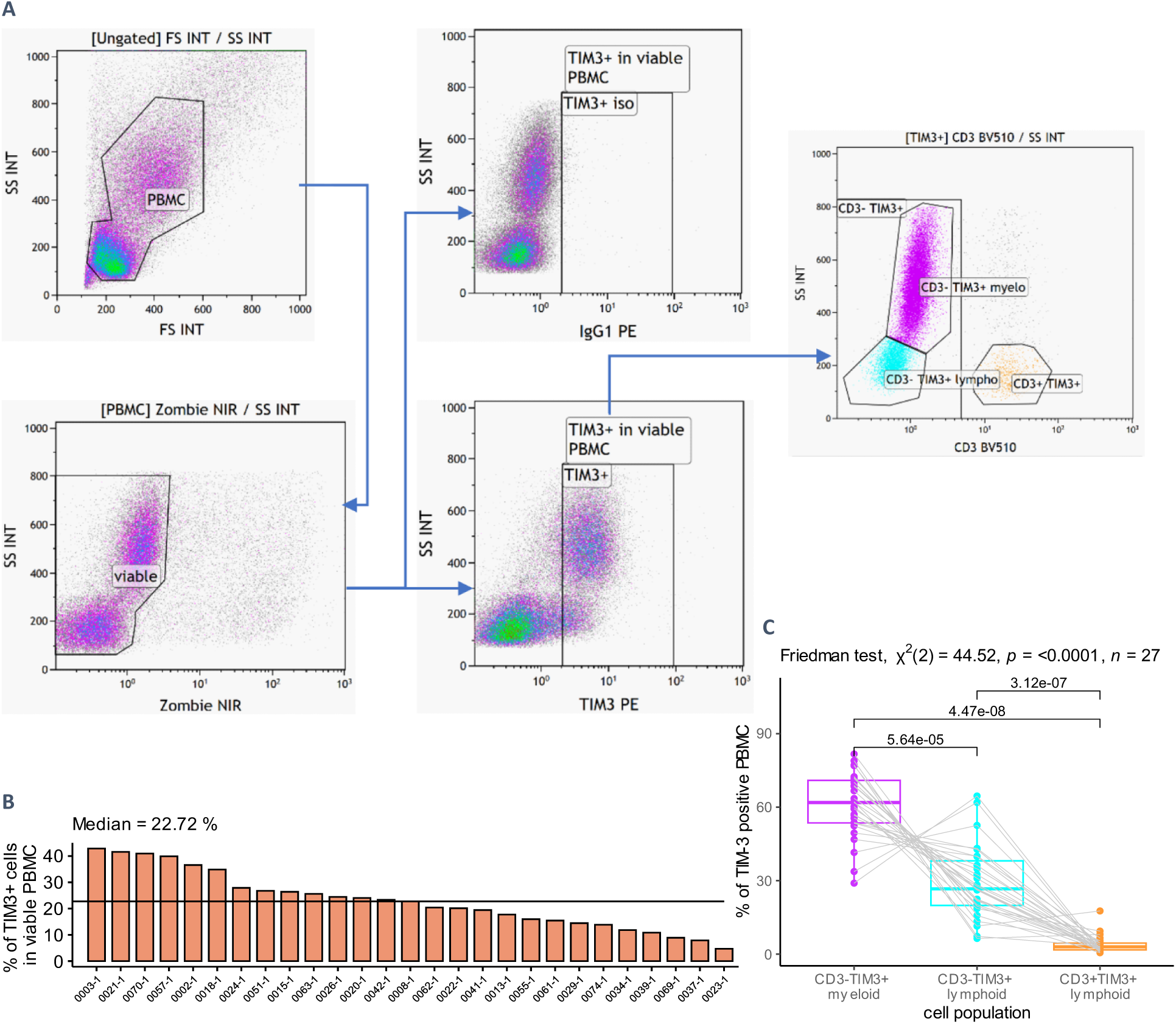
Flow cytometry quantification of TIM-3-positive (TIM3+) cells in PBMC of ccRCC patients (Colcheckpoint cohort, n = 27). A. Gating strategy; we considered CD3-SSC-A^high^ cells as myeloid cells, CD3+SSC-A^low^ as T cells and CD3-SSC-A^low^ as non-T lymphoid cells. B. Percentage of TIM3+ cells within PBMC of Colcheckpoint participants. C. Proportion of CD3- myeloid, CD3- lymphoid and CD3+ lymphoid cells within TIM3+ cells in PBMC of Colcheckpoint participants

HAVCR2+ADAM+ enriched monocytes clusters were common to paired tumors and PBMC in a dataset of 3 ccRCC patients ^20^. Consistent with our flow cytometry data, HAVCR2 expression was mainly found on monocytes and NK clusters in PBMC (**Supp.Fig.13**).

Principal component analysis (PCA) on the top 200 most variable genes showed a clear separation of double-positive and non-double-positive pseudobulk samples, suggesting a distinct transcriptomic state for HAVCR2+ADAM+ myeloid cells (**Supp.Fig.13**).

Among top DEG, the putative pro-tumor markers TREM2 and APOE were found increased in HAVCR2+ADAM+ myeloid cells (Khantakova, Brioschi, et Molgora 2022; Bancaro et al. 2023), while NLRP3 was decreased, which may be linked to decreased APC functionality (Dixon et al. 2021). GSEA suggested a downregulation of proteins synthesis in HAVCR2+ADAM+ myeloid cells (**Supp.Fig.14**), as wells as cellular respiration (oxidative phosphorylation).

These results favor myeloid cells as major source of sTIM3 in mccRCC. sTIM-3 could reflect the presence of dysfunctional HAVCR2+ADAM+ myeloid cells, with globally decreased anabolic activity ^23^.

#### C. PBMC cytometry of ccRCC patients

We quantified TIM-3-positive (TIM3+) cells in PBMC from mccRCC patients (**Fig.5.A**), their median proportion was 22.7% (IQR 14.9% to 27.3%) (**Fig.5.B**). CD3-TIM3+ myeloid cells were the most common type of TIM3+ PBMC (median 61.9% of TIM3+ PBMC), followed by CD3-TIM3+ lymphoid cells. Surprisingly, T cells (CD3+TIM3+) accounted for a minority of TIM3+ PBMC (**Fig.5.C**). Given their intermediate CD4 positivity (**Supp.Fig.15**) and that their proportion among TIM3+ PBMC was the same as CD14+TIM3+ PBMC on a separate panel (**Supp.Fig.16**), we hypothesized that most CD3-TIM3+ myeloid cells had a monocytic origin. CD3-TIM3+ lymphoid PBMC were most likely NK cells, given their CD3-CD8low phenotype and that B cells (quantified with CD19 on another panel) were not found in TIM3+ PBMC. These results are consistent with data from the literature on peripheral blood cells from healthy donors, showing TIM-3 detection on a high proportion of monocytes, on a moderate proportion of NK cells, and low or absent detection on T cells and granulocytes (Kikushige et al. 2010; Möller-Hackbarth et al. 2013).

#### D. Lymphocytes depletion in TC-1 syngeneic mouse model

Serum sTIM-3 increase upon antitumor vaccination was found at earlier time-points in a second experiment with the TC-1 mouse model (**Supp.Fig.20.A**). Given our initial hypothesis of sTIM-3 release by T cells following vaccination, control vaccinated mice were compared to mice vaccinated and depleted in CD8+ T cells with an anti-CD8α depleting monoclonal antibody. Effective depletion was confirmed by the quasi-disappearance of T cells in tumors from anti-CD8-treated mice analyzed by flow cytometry (not shown). However sTIM-3 levels were not significantly modified by the depletion (**Supp.Fig.20.B**). The timing of sTIM-3 increase and its sustainability in spite of CD8+ T cells elimination favor the hypothesis of the early release of sTIM-3 by innate immune cells, possibly myeloid cells.

## Discussion

Plasma sTIM-3 is a promising biomarker associated with ICI efficacy in mccRCC, with differential effects on nivolumab versus N+I and, independent from common clinical or inflammatory markers. An association with survival of patients under current anti-PD-1 + TKI regimens is also plausible and will be investigated in future research. If a differential effect on N+I compared to anti-PD-1 + TKI is confirmed, this could make sTIM-3 a theranostic biomarker. Defining a sTIM-3-high population for which the addition of the anti-CTLA-4 is needed to overcome the resistance to anti-PD-1 would facilitate the indication of N+I, for which the benefit-risk balance is delicate. sTIM-3 is also promising for other cancers where anti-PD-1 monotherapies are challenged by combinations, without strong decisional criteria (e.g. melanoma, lung cancer).

TIM-3 is known for its inhibitory role at the membrane of T cells, its shedding into sTIM-3 by activated CD8+ T cells was reported ^14^. Unexpectedly, our study suggests myeloid cells as major source of sTIM- 3 in mccRCC. TIM-3 was found on tumor-associated macrophages (TAM) and associated with PFS, in another cohort of ccRCC, and induced on monocytes co-cultured with RCC cell-lines. However, the patients were not treated with ICI and monocytes were obtained from healthy volunteers ^24^. LPS-activated monocytes from healthy donors and myeloid leukemia cells shed sTIM-3 upon activation of ADAMs ^13,25^. We show in mccRCC that myeloid cells represent the majority of TIM-3-positive PBMC and the most enriched lineage in HAVCR2-expressing cells within the TME. A low proportion of TIM-3- positive lymphoid cells in cytometry could be due to shedding of the marker ^13,14^. Nevertheless, scRNAseq shows more co-expression of ADAM10/17 in myeloid cells, thus, a bias on membrane TIM-3 detection is more likely to occur for them. TIM-3 was detected at the apical border of some healthy and cancerous proximal tubule cells in IHC (**Fig.5**). One hypothesis is that sTIM-3 undergoes glomerular filtration with re-uptake of the antigen by these cells^26^. In our analysis of Chevrier et al. mass cytometry dataset (**Supp.Fig.15**), TIM-3 appeared fairly expressed by immune cells but with null median signal intensities for non-immune cells in most samples (**Supp.Fig.15<u>.B&C</u>**). Finally, granulocytes are quasi-absent from PBMC and from the scRNAseq datasets but their role as source of sTIM-3 cannot be excluded.

A secreted sTIM-3 isoform comes from alternatively splicing of murine TIM-3 mRNA ^27^. Nonetheless, mouse embryonic fibroblasts shed transfected human TIM-3 and murine TIM-2 (TIM family) via ADAM10/17, supporting the relevance of studying sTIM-3 in this species ^13,28^.

Regarding the biological meaning of sTIM-3 in mccRCC patients, there are two (non-exclusive) possibilities: sTIM-3 could have *per se* a pro-tumor effect ^25,29^. On the other hand, plasma sTIM-3 could be a marker of pro-tumor processes or cell types, such as ADAM10/17 activity in the TME ^30,31^. We show that HAVCR2+ADAM+ myeloid cells have an increased transcription of TREM2, which is proposed as a pro-tumor macrophage marker ^32^. Vanmeerbeek et al. have identified TAM co-expressing TIM-3 and VSIR, associated with resistance to ICI, HAVCR2 was upregulated in myeloid cells from ICI-unresponsive patients^33^. The release of sTIM-3 may also reflect a deleterious state of over-activation of myeloid APC, which are required to prime T cells and sustain the response to ICI ^34–36^. GSEA suggest that HAVCR2+ADAM+ myeloid APC have globally impaired transcriptional and translational activities (Supp.Fig.13) ^37,23^.

Understanding the biological meaning of plasma sTIM-3 will be crucial for TIM-3-targeted therapies. According to the relevant hypothesis, strategies to treat sTIM-3 high patients would be either to block the putative deleterious action of sTIM-3, to inhibit ADAMs’ activities, to deplete TIM-3+ pro-tumor myeloid cells or to restore the function of paralyzed APC.

## Conclusion

- Plasma sTIM-3 is elevated in mccRCC and independent from clinical and inflammatory prognostic markers. It is a promising blood-based biomarker associated with ICI efficacy in this setting.
- The immune myeloid compartment is the predominant candidate as source of plasma sTIM-3 in mccRCC.
- Further functional studies will precise the biological role of sTIM-3 in mccRCC and ensuing therapeutic targets.

## Supplementary figures

**Supp.Fig.1:**
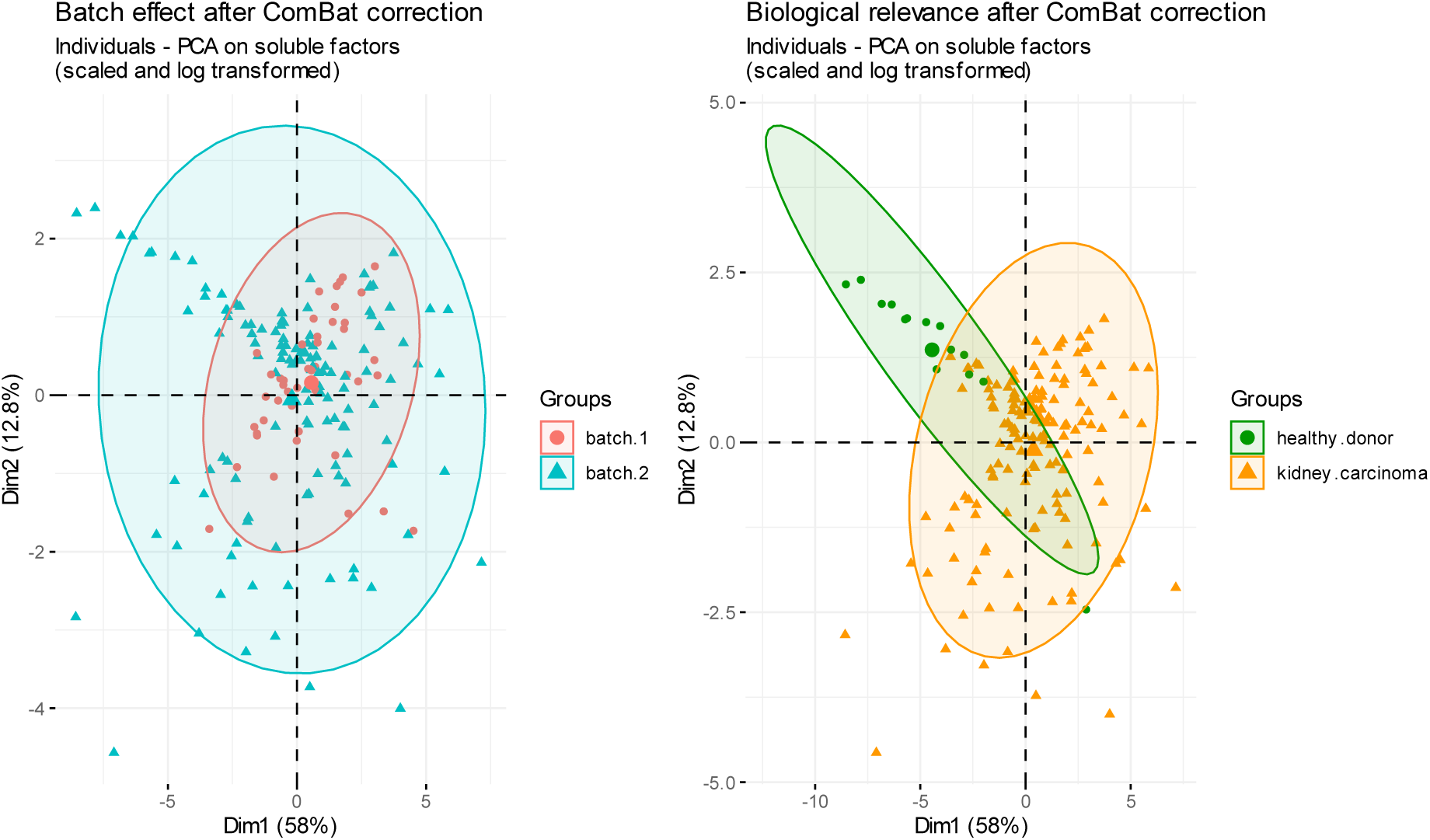
PCA of batch-corrected soluble factors values for participants of the BIONIKK study. In order to minimize the influence of a potential batch effect of the Luminex quantification method, sTIM3 values were batch-corrected through the ComBat() function from the sva R package (v. 3.50.0) prior to classification of individuals for comparisons between the nivolumab and N+I arms of the BIONIKK cohort. This method enabled to successfully limit technical variability between batches, while preserving biological significance. As a control, healthy donors were clearly distinguished from ccRCC patients after dimensional reduction of the data of other soluble proteins quantified along sTIM-3 in these individuals

**Supp.Fig.2:**
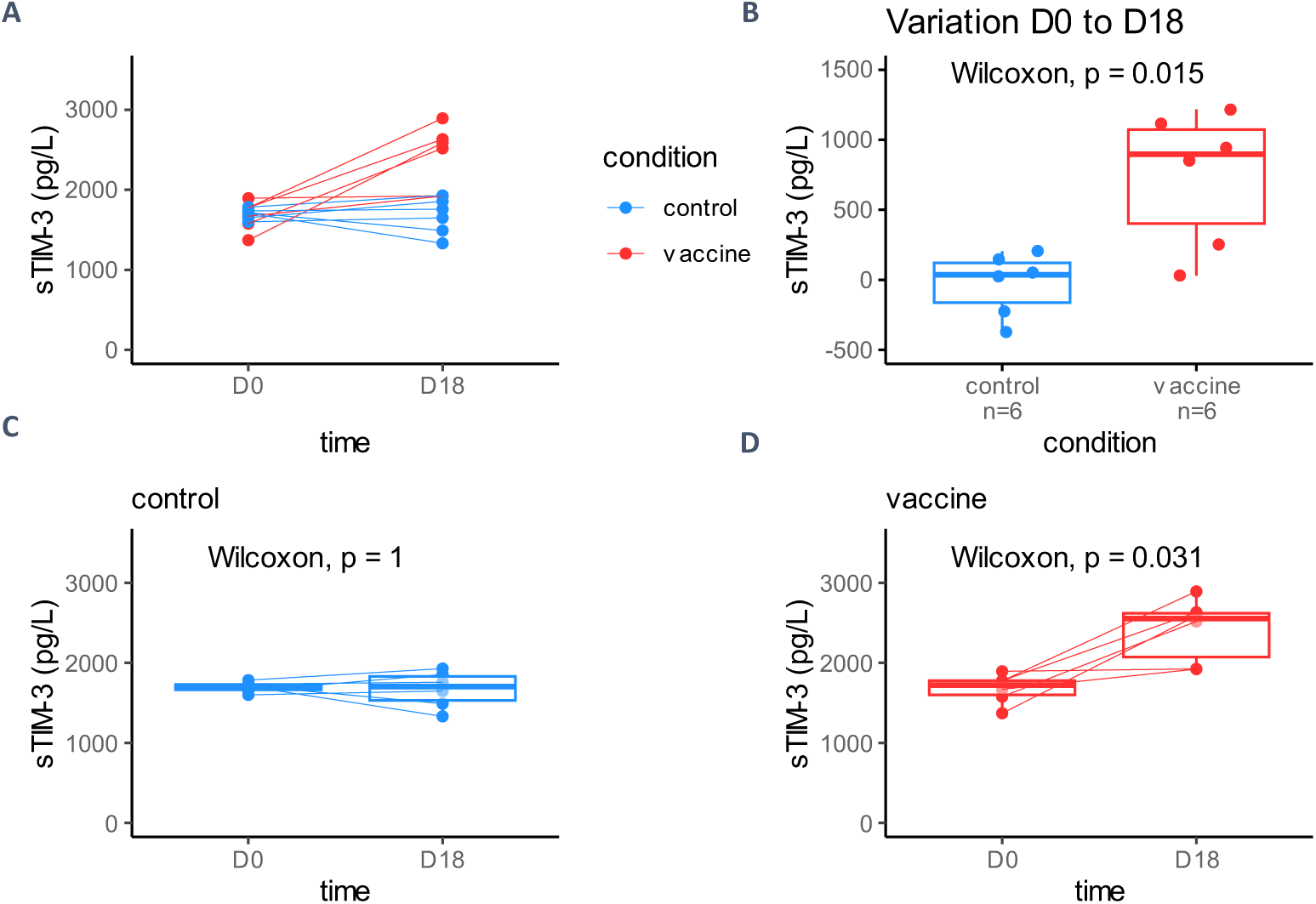
sTIM-3 plasma levels in TC-1 syngeneic mice immunized with an antitumor vaccine. **A.** TC-1 syngeneic mice were inoculated (intra-jugal, 50,000 cells/50 µL) with TC-1 tumor cells (murine pulmonary epithelial line immortalized by the E6 and E7 proteins from human papillomavirus 16 – HPV-16) at day 0 (D0) and then vaccinated (vaccine group, injection at D7 and D14) or not (control group), plasma sTIM-3 was measured at D0 and D18, n = 6 individuals for both groups. **A.** All measures. **B.** Comparison of sTIM-3 variation (D18 values minus D0 value for each individual) between control and vaccine group (Wilcoxon rank sum test). **C & D.** Measure of sTIM-3 levels over time within control and vaccine groups respectively (Wilcoxon matched pairs test).

**Supp.Fig.3:**
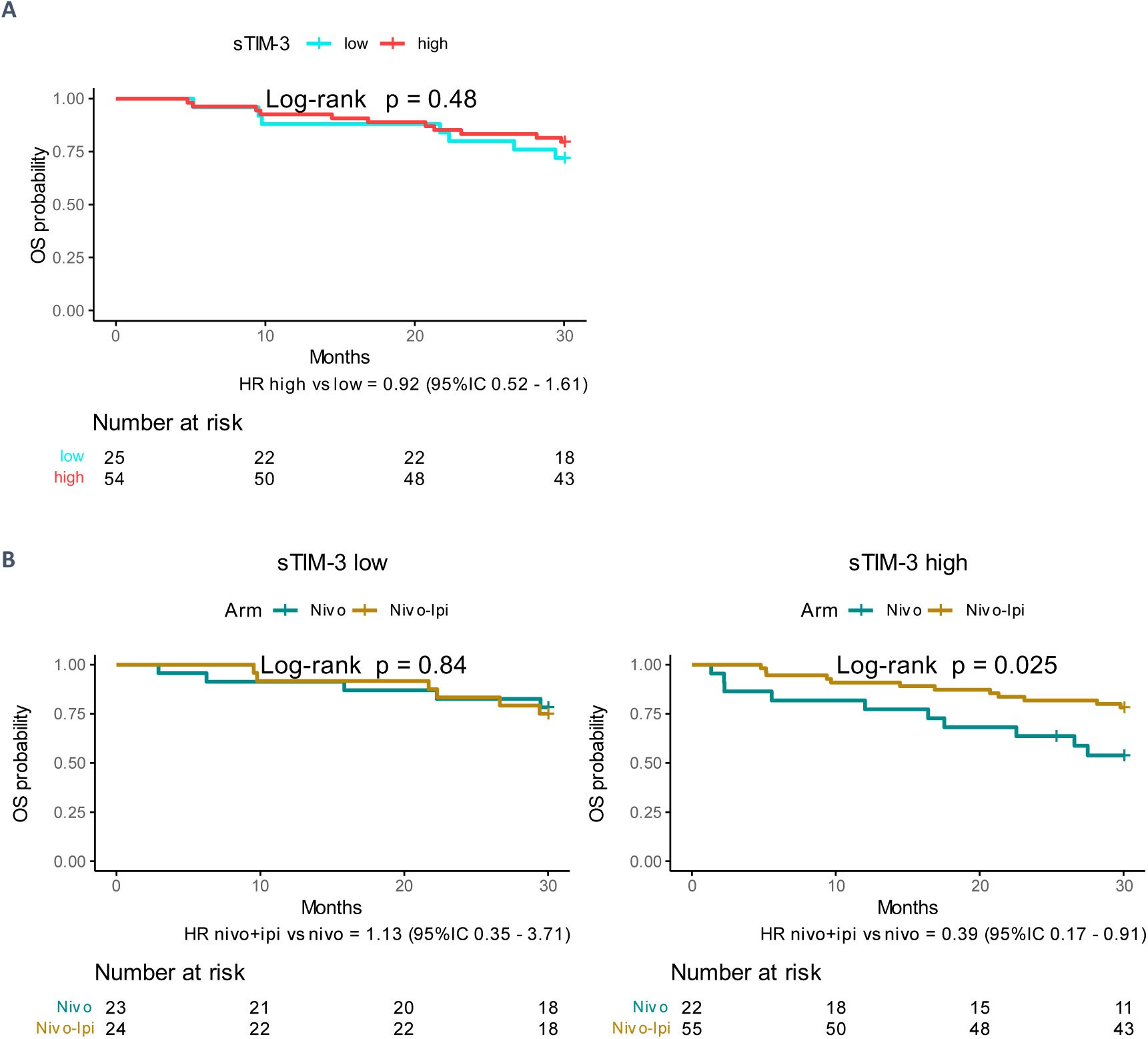
Association of plasma sTIM-3 and OS in patient from the BIONIKK trial treated with nivolumab + ipilimumab combination. **A.** OS in sTIM-3 high vs sTIM-3 low mccRCC patients treated with nivolumab + ipilimumab and **B.** Comparison of OS between nivolumab + ipilimumab and nivolumab monotherapy within each sTIM-3 strata.

**Supp.Fig.4:**
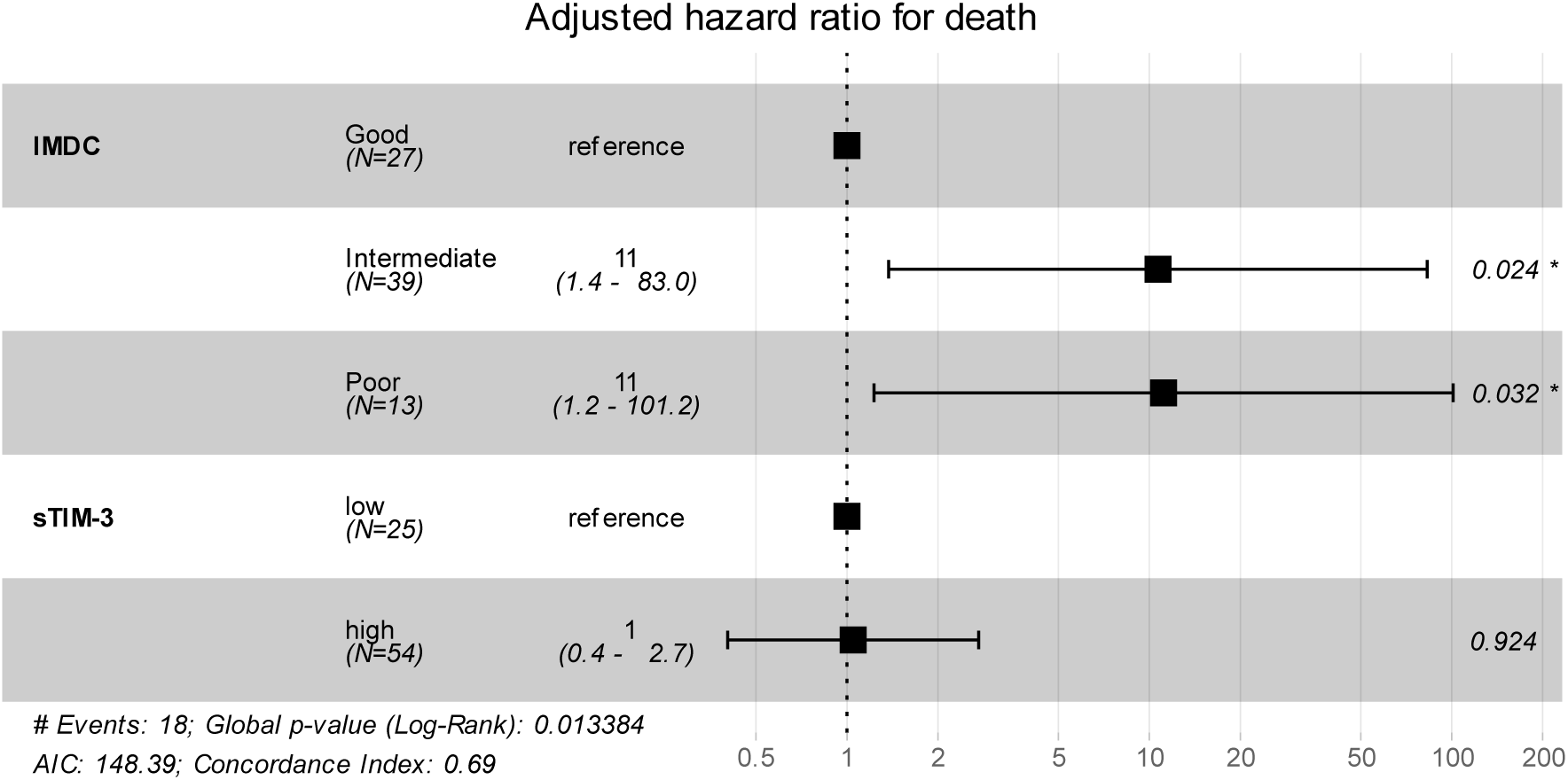
Multivariable Cox regression model for OS, adjusting for IMDC in BIONIKK nivolumab + ipilimumab participants.

**Supp.Fig.5:**
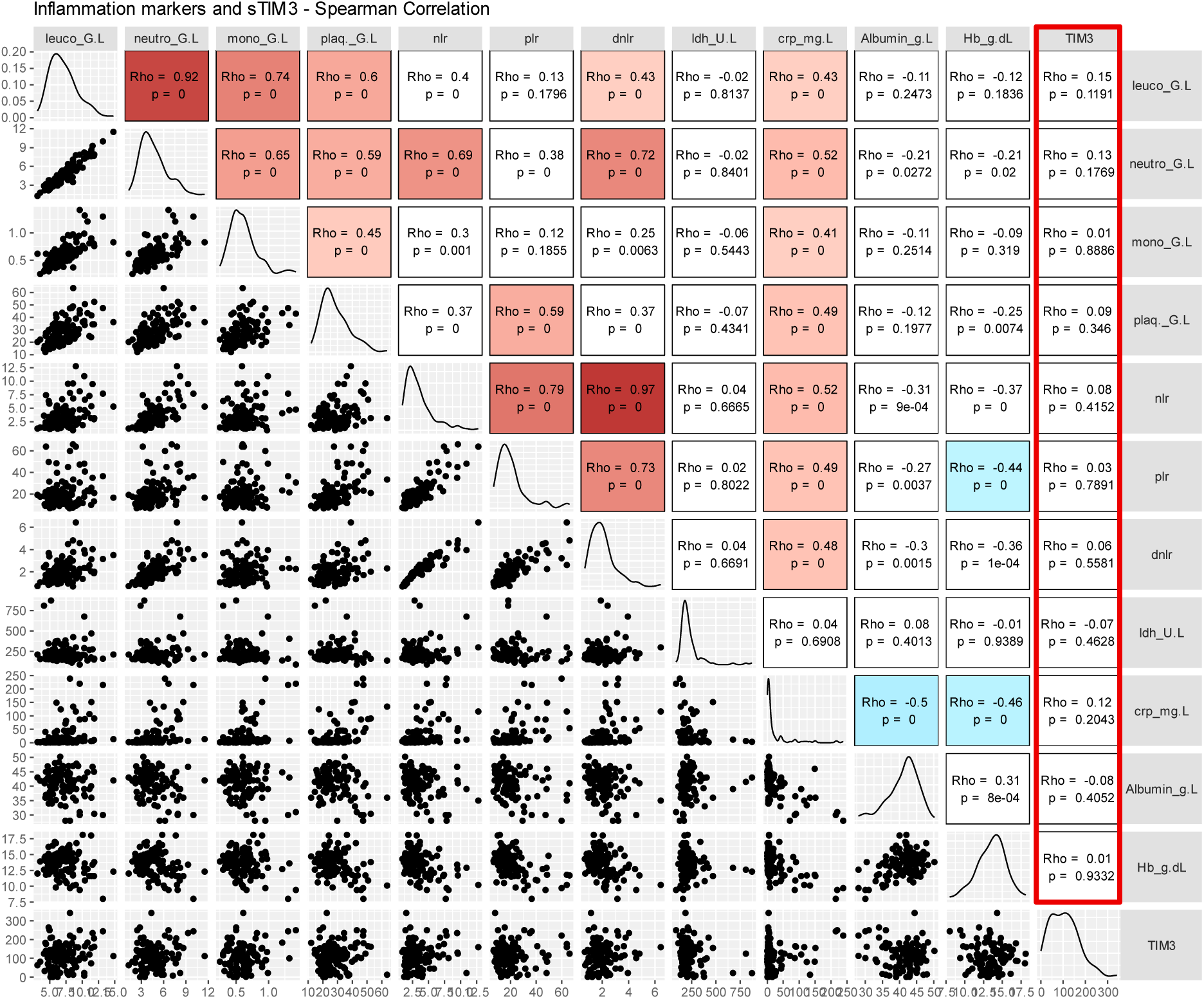
Spearman correlation matrix between classical inflammatory markers and sTIM-3.

**Supp.Fig.6:**
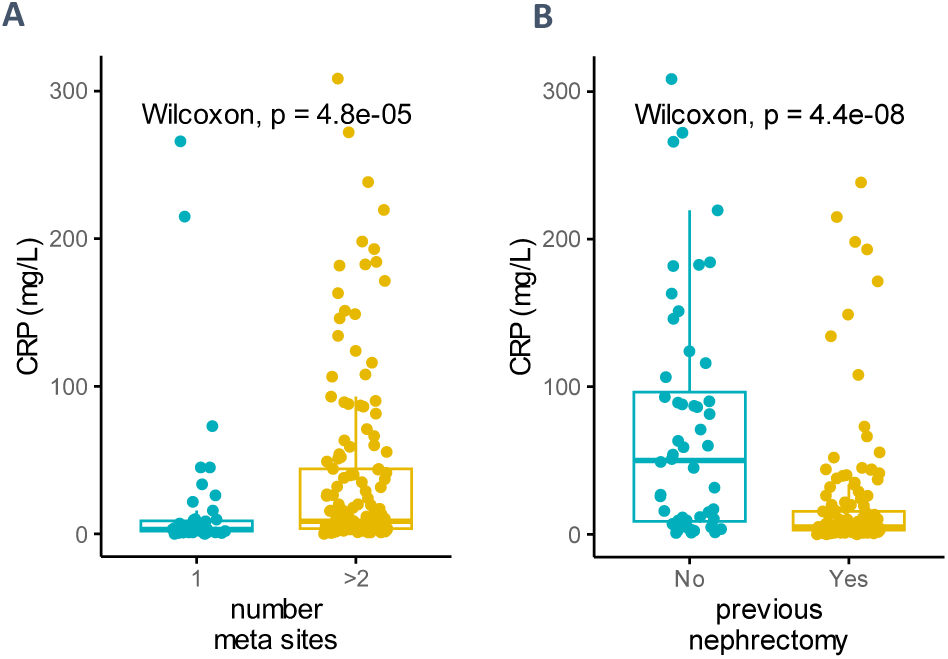
Comparisons of serum CRP in BIONIKK participant. **A.** 1 versus ≥2 metastatic sites. **B.** Primary tumor resected (“previous nephrectomy yes”) versus not resected.

**Supp.Fig.7:**
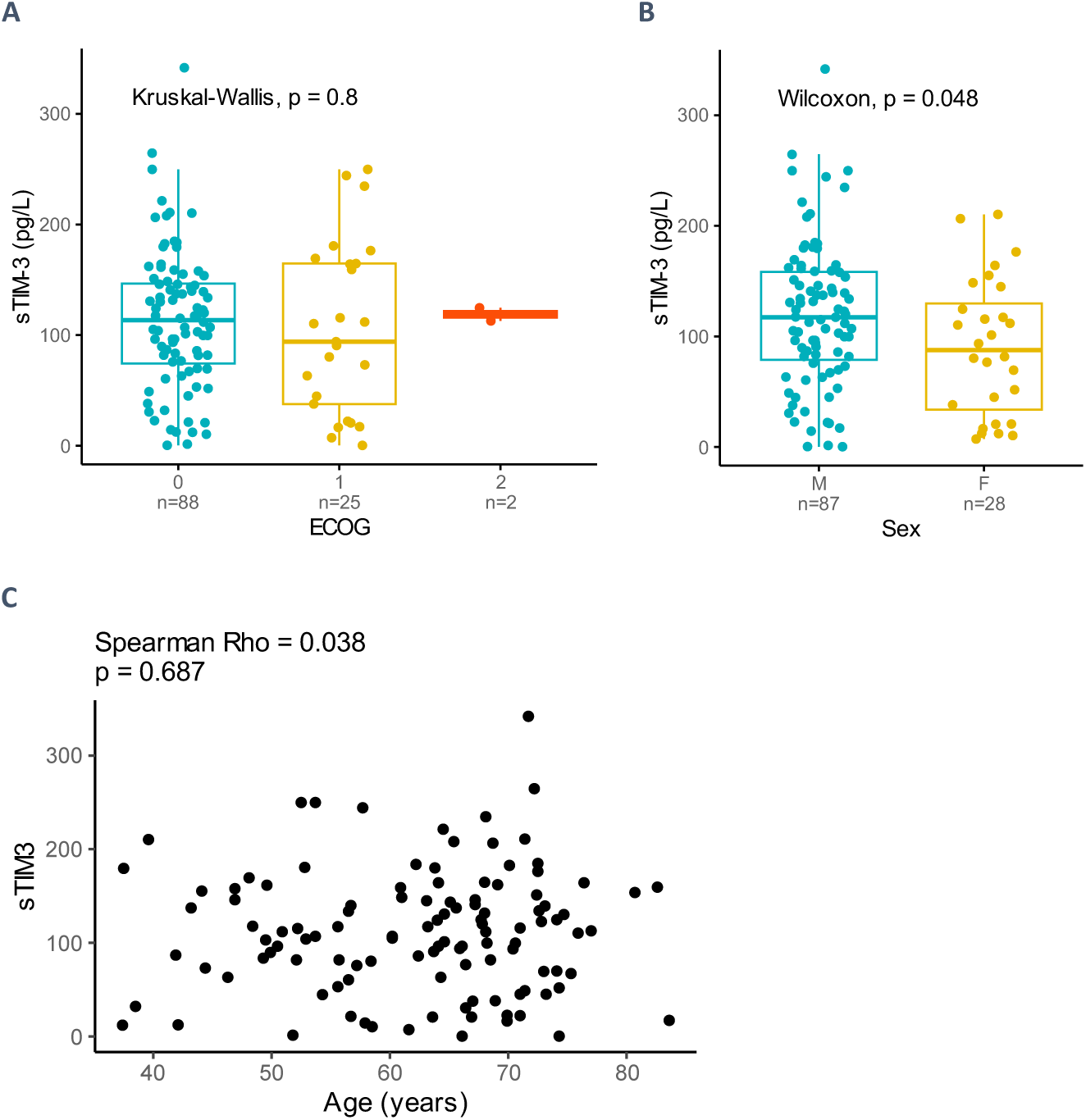
Association of sTIM-3 with clinical variables in BIONIKK participants: **A.** sTIM-3 versus ECOG performance status. **B.** sTIM-3 versus sex. **C.** sTIM-3 versus age.

**Supp.Fig.8:**
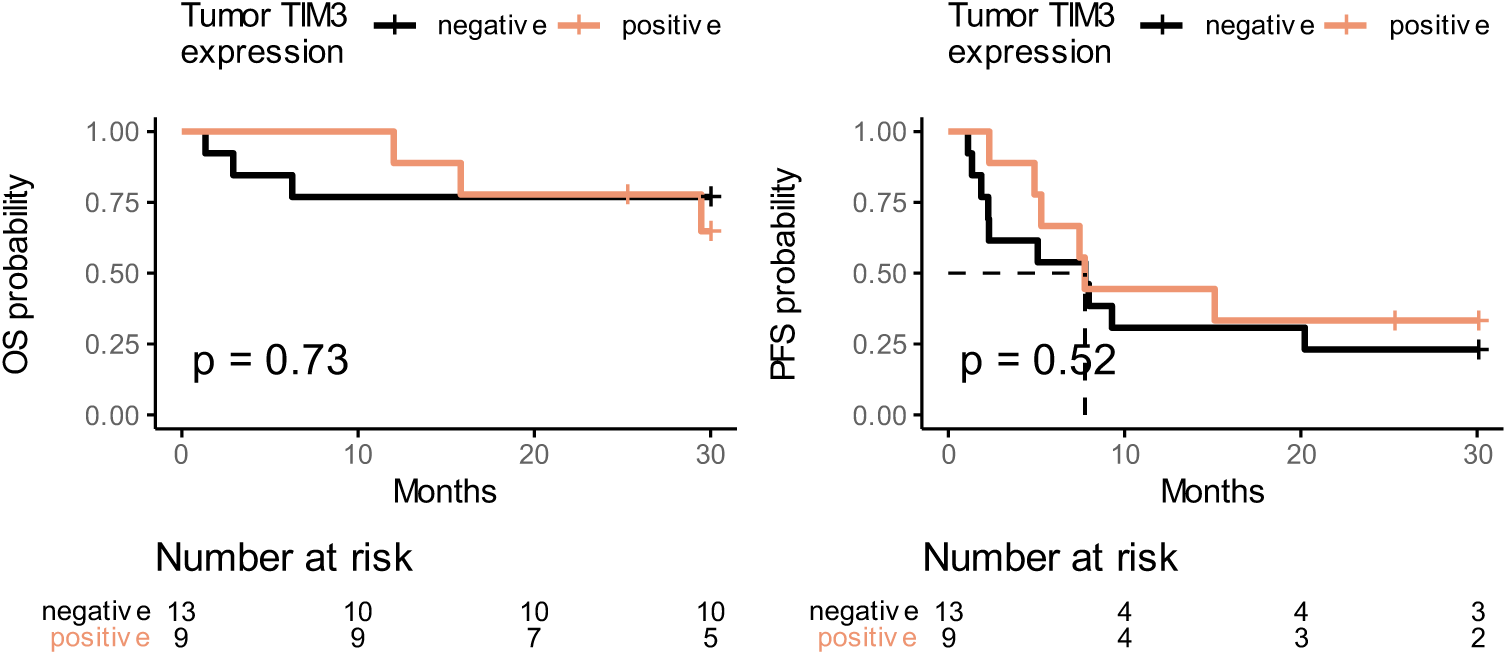
OS (left) and PFS (right) in the BIONIKK IHC subgroup of participants treated with Nivolumab monotherapy stratified according to tumor TIM-3 staining.

**Supp.Fig.9:**
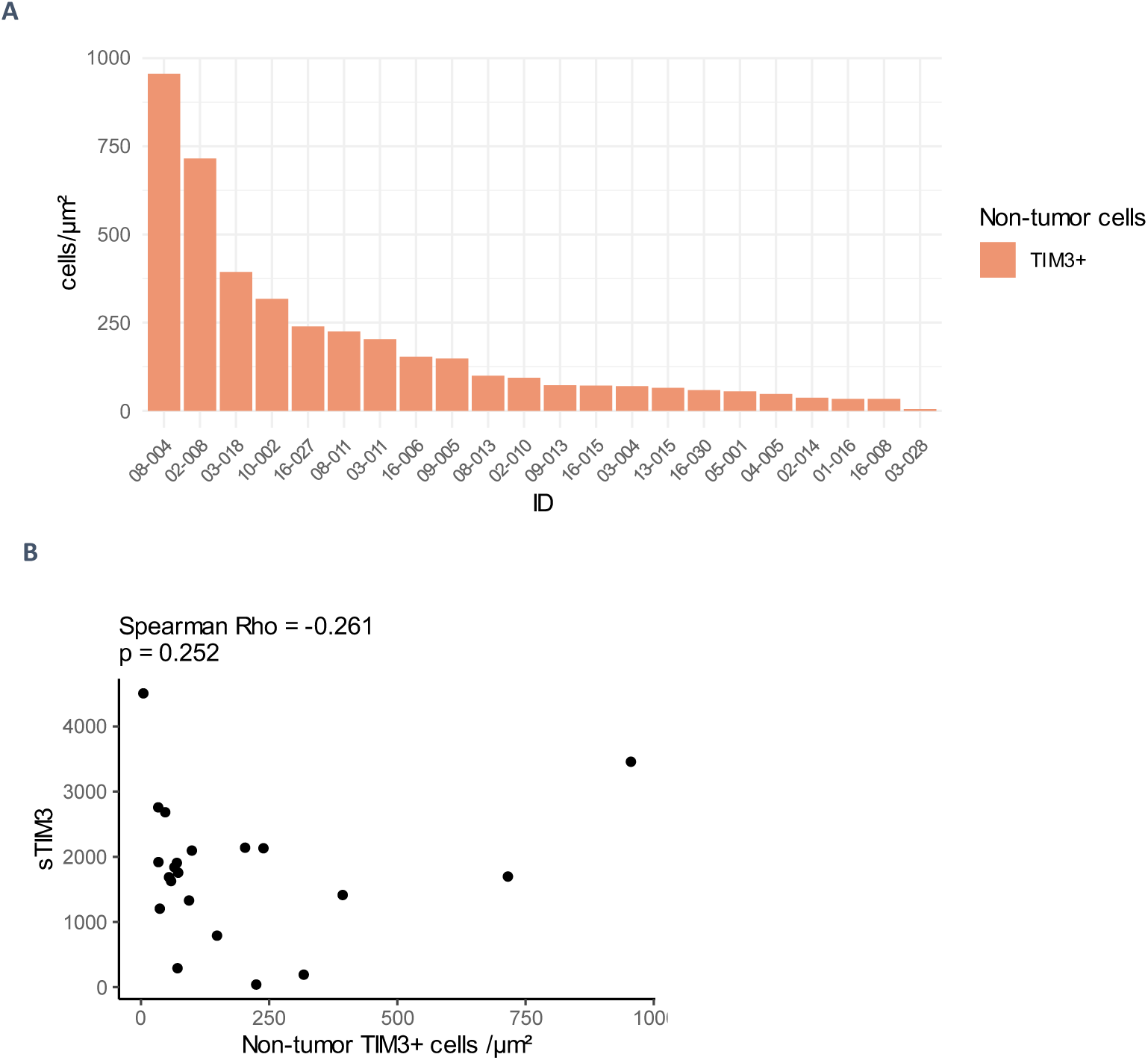
Membrane TIM-3 on non-tumor cells in IHC. **A.** Density of non-tumor TIM-3 positive cells assessed by IHC in tumors of the BIONIKK patients IHC subset. **B.** Correlation plot between non-tumor TIM-3 positive cells density and sTIM-3 plasmatic levels

**Supp.Fig.10:**
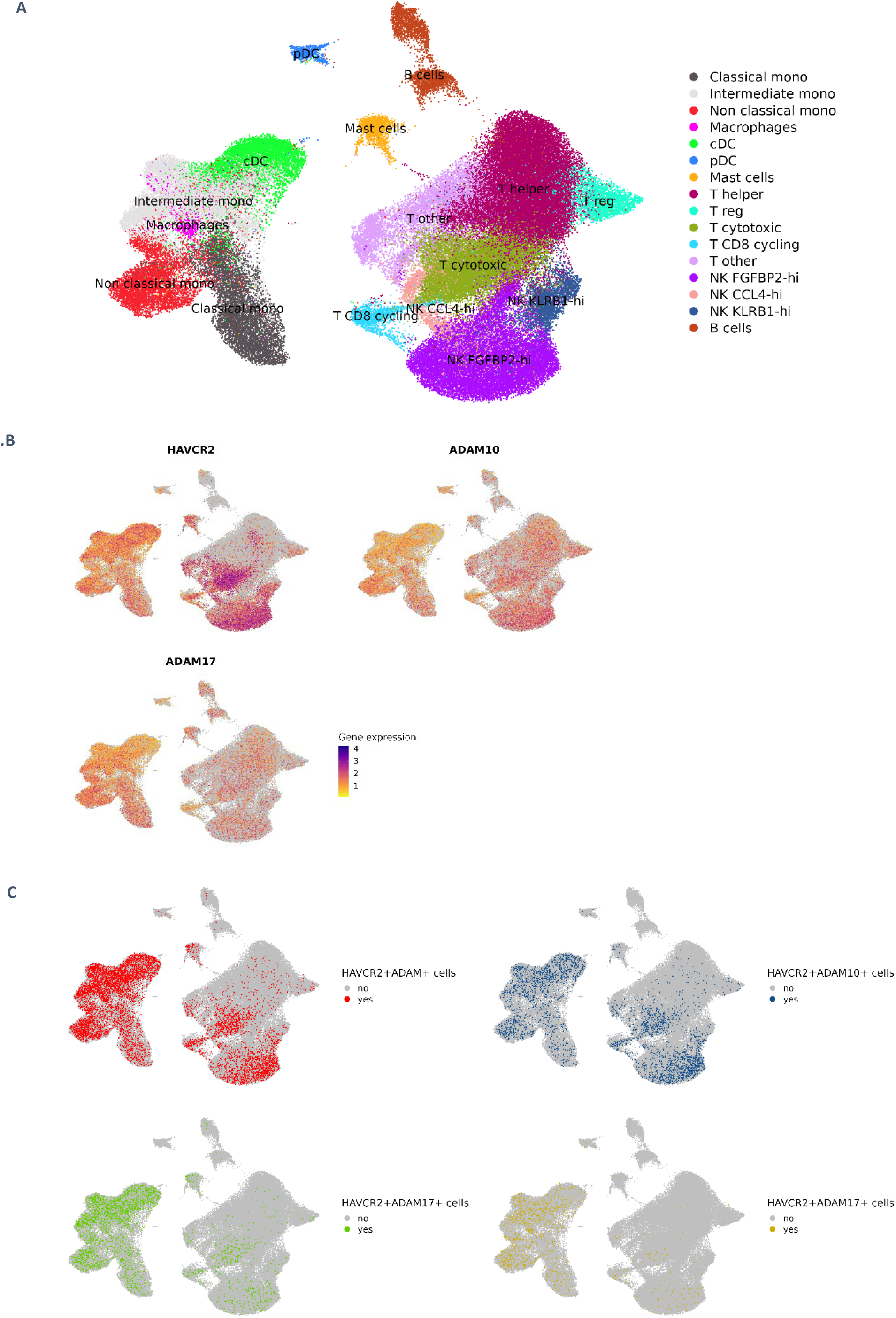
UMAPs of CD45 positive cells from the scRNAseq dataset of Obradovic et al. **A.** Detailed clusters. **B.** Gene expression levels for HAVCR2, ADAM10 and ADAM17. **C.** Detail of HAVCR2 and ADAM10/17 co-expression phenotypes.

**Supp.Fig.11:**
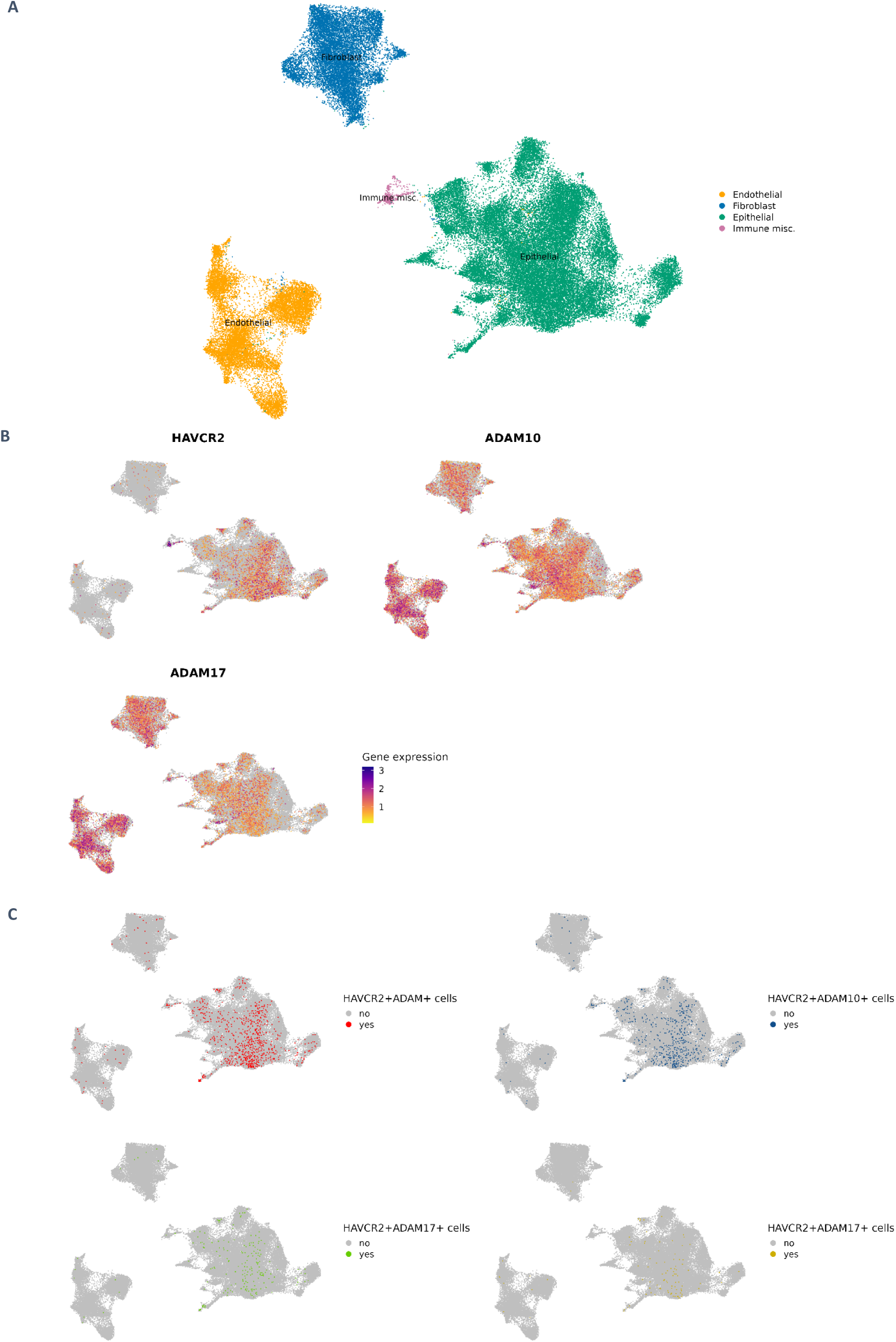
UMAPs of CD45 positive cells from the scRNAseq dataset of Obradovic et al. **A.** Detailed clusters. **B.** Gene expression levels for HAVCR2, ADAM10 and ADAM17. **C.** Detail of HAVCR2 and ADAM 10/17 co-expression phenotypes.

**Supp.Fig.12:**
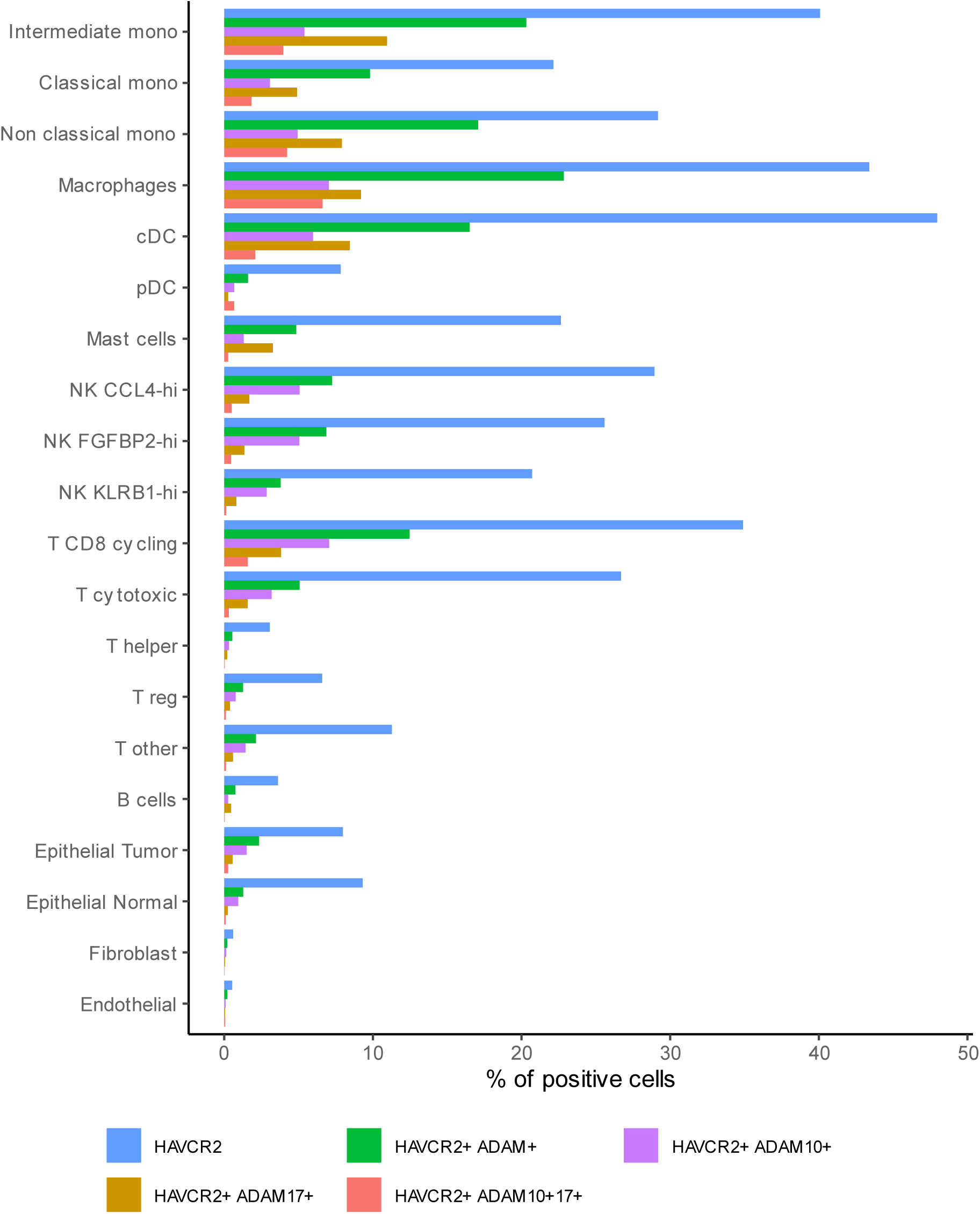
Percentage of cells expressing HAVCR2 or co-expressing HAVCR2, ADAM10, ADAM17, within each cluster in the scRNAseq dataset of Obradovic et al.

**Supp.Fig.12:**
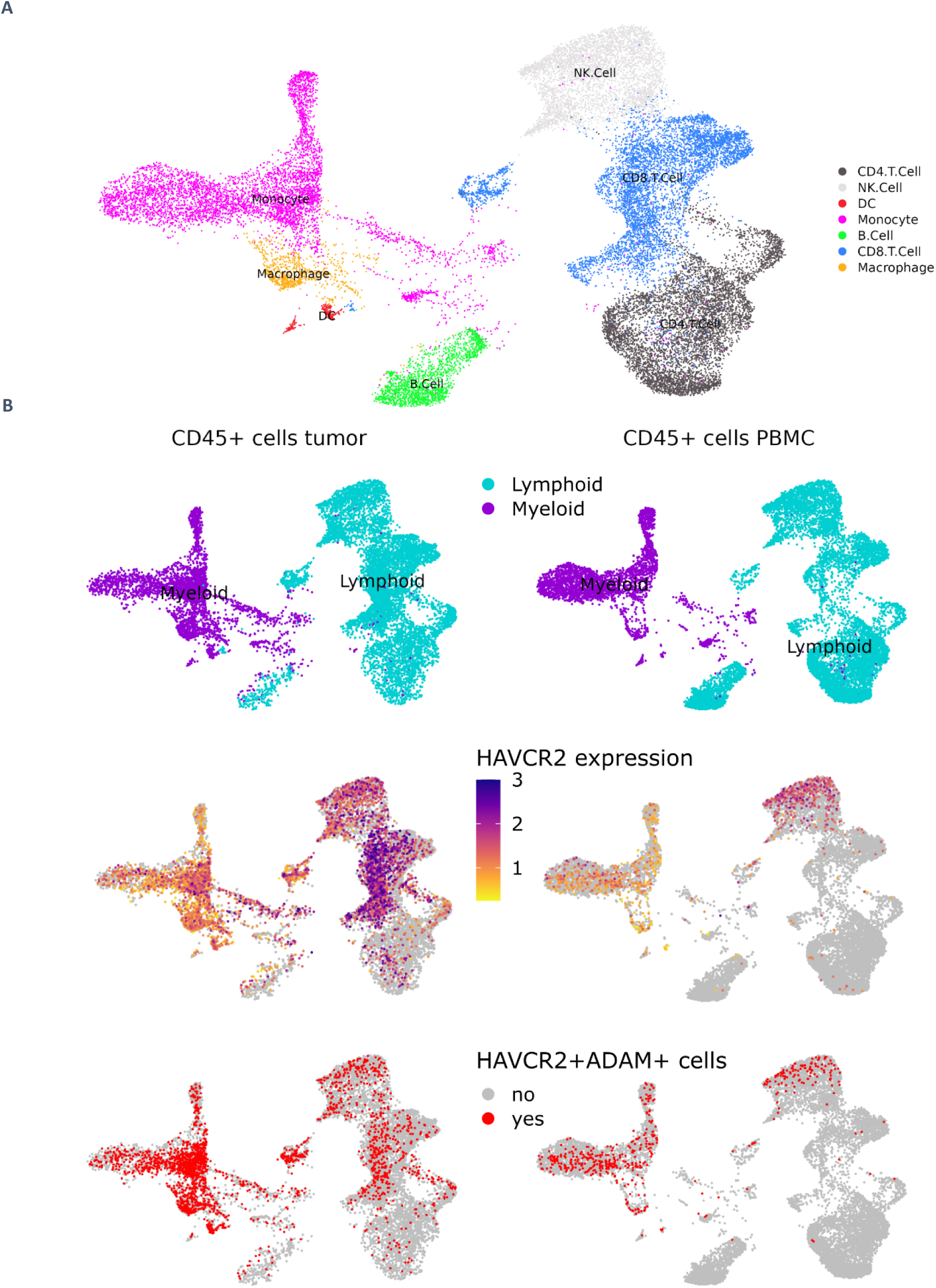
UMAPs of CD45 positive cells from 3 ccRCC patients in the scRNAseq dataset of Borcherding et al. **A.** Detailed clusters **B.** Comparison of immune cells from matched tumor and PBMC samples. upper section: UMAP of cell lineages. middle section: HAVCR2 expression. lower section: HAVCR2+ADAM+ double-positive cells repartition

**Supp.Fig.13:**
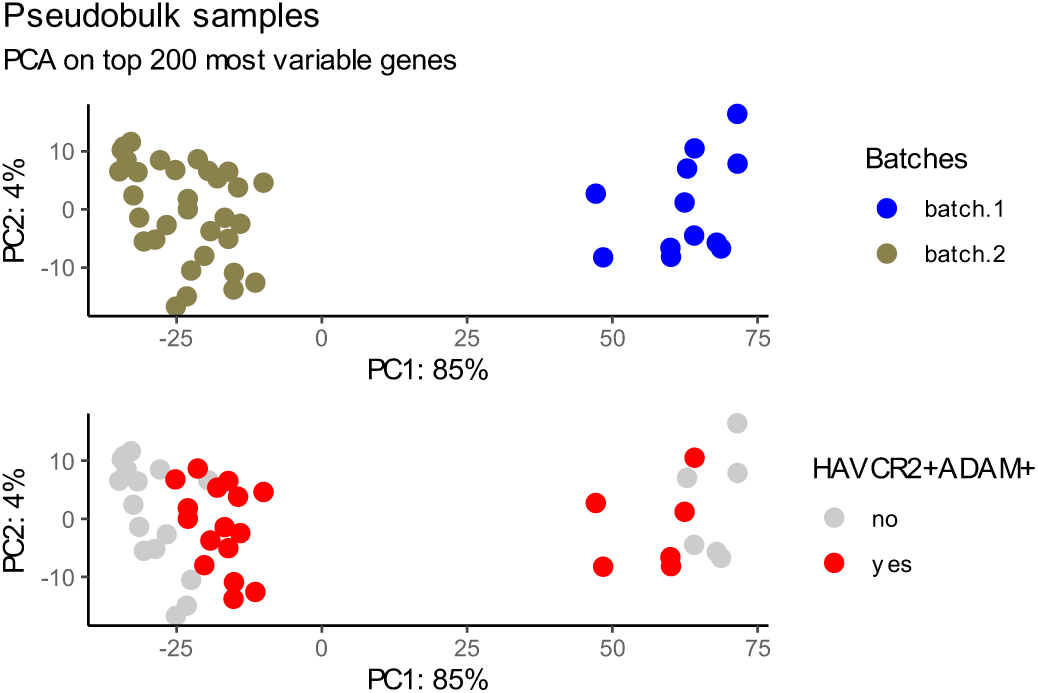
Pseudobulk samples of HAVCR2+ADAM+ double-positive and non-double-positive myeloid cells from Obradovic et al. scRNAseq dataset. Projection of samples on the first two axes of PCA on the top 200 most variable genes. Transcripts counts cells were aggregated in pseudobulk samples to compare HAVCR2+ADAM+ myeloid cells to other myeloid cells. Top panel: visualization of the batch effect. Bottom panel: segregation of double-positive and non-double-positive pseudobulk samples within each batch.

**Supp.Fig.14:**
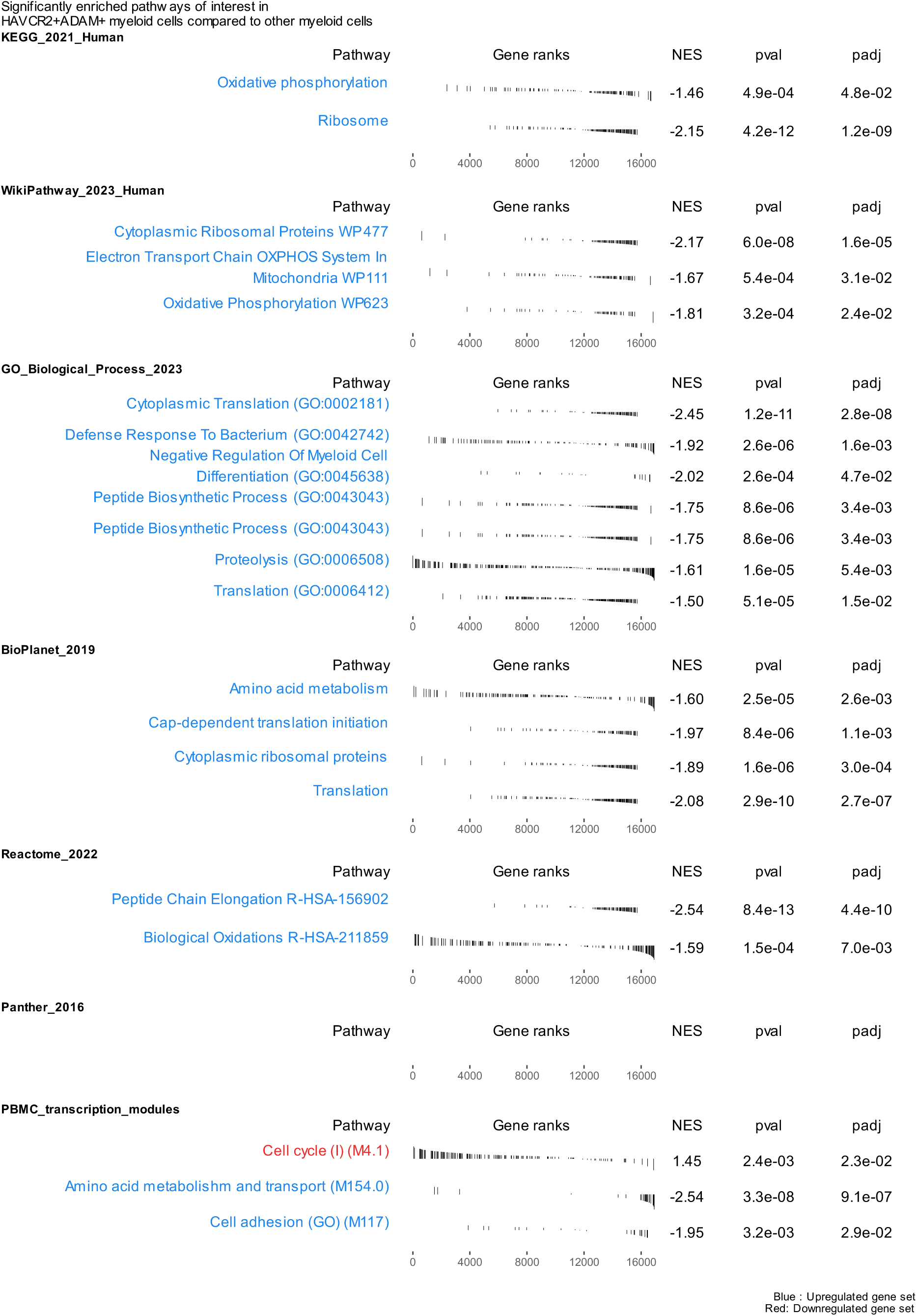
Pathways of immunological interest with significant enrichment after GSEA on differentially expressed genes between HAVCR2+ADAM+ myeloid cells and other myeloid cells from Obradovic et al. NES: normalized enrichments score. Padj: adjusted p-value.

**Supp.Fig.15:**
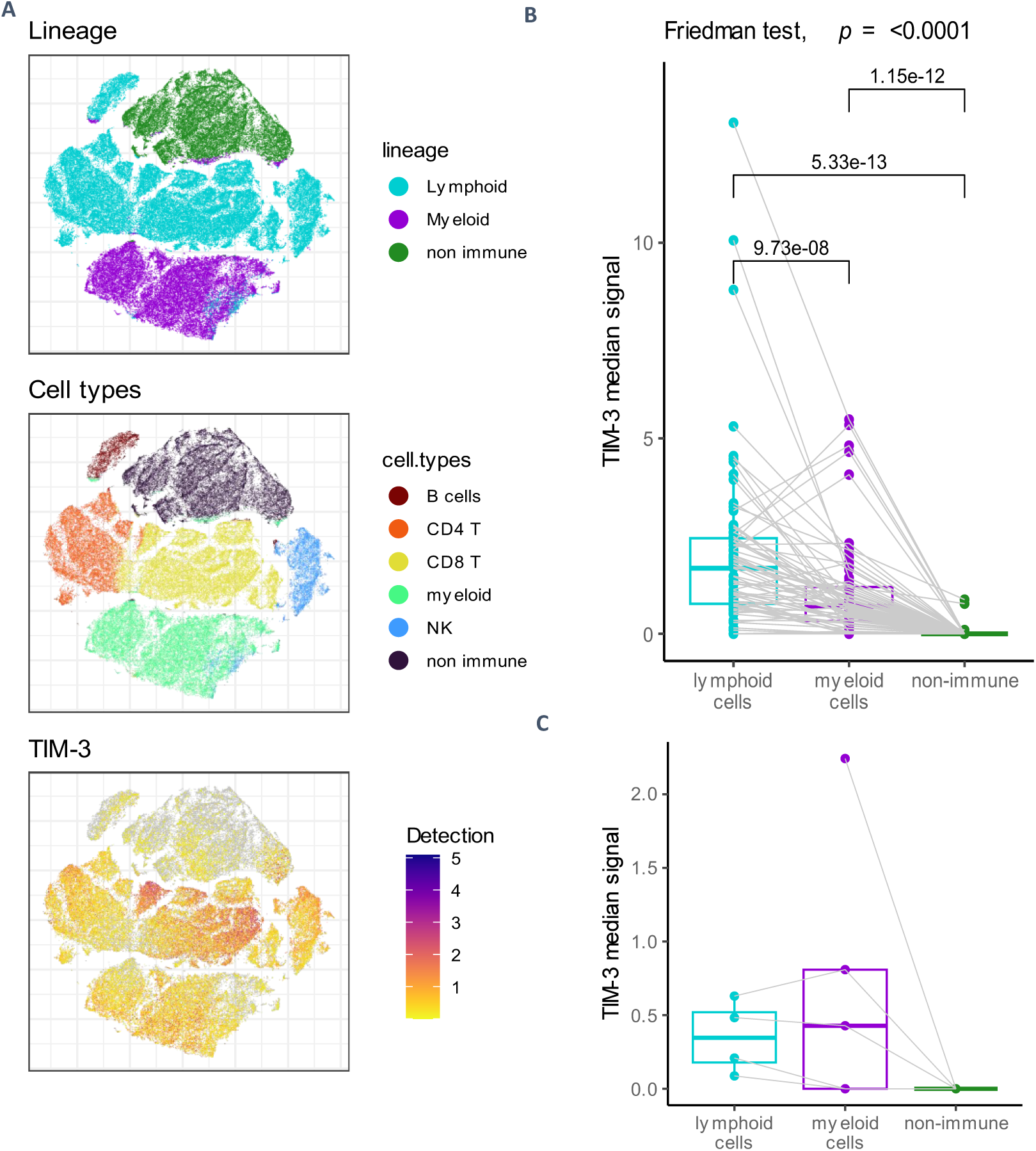
Mass cytometry quantification of TIM-3-expression cells in the dataset of Chevrier et al. of 72 ccRCC tumors and 5 healthy kidney samples. Publicly available mass cytometry “T cell panel” data from Chevrier et al. was accessed at https://premium.cytobank.org/cytobank/projects/875 ^38^. .fcs files correspond to debarcoded samples of viable cells, filtered out for gadolinium contamination and cells doublets. The data is bead-based normalized. We applied the standard arcsinh transformation with a cofactor of 5 on the value of all channels for clustering and visualization purposes. A compensation matrix was calculated with the CATALYST R package (v. 1.18.1) according to the procedure and single-stained beads reference assays reported by Chevrier et al.^39^. Given the absence of significant changes after applying compensation (**Supp.**Fig.17), the data was left uncompensated. Annotation of cells by broad lineages and immune cell types was performed by unsupervised k-means clustering and examination of the expression profile of consensus markers. Contrary to the procedure used by Chevrier et al., markers such as TIM-3 and other immune checkpoint were not included in the matrix used to cluster cells, to avoid the artificial construction of TIM-3-high clusters. The data was scaled prior to k-means clustering and a ponderation was used with higher weights attributed to major markers such as CD45 and CD3. The expression of TIM-3 and other markers on single cells was visualized on dimensionality reduction 2-D projections generated with the t-SNE algorithm, after subsampling the whole dataset to a maximum of 1500 cells per sample for convenience. For statistical comparisons, the median signal intensity of TIM-3 was calculated on untransformed values. **A.** tSNE representation of cell clusters and TIM-3 expression (asinh transformed values) after subsampling of the dataset. **B.** Median signal intensity (untransformed) for TIM-3 in lymphoid, myeloid and non-immune cells of the ccRCC samples. P-values are given on brackets for paired Wilcoxon tests between each lineage. **C.** Median signal intensity (untransformed) for TIM-3 in lymphoid, myeloid and non-immune cells of the healthy donors’ samples.

**Supp.Fig.16:**
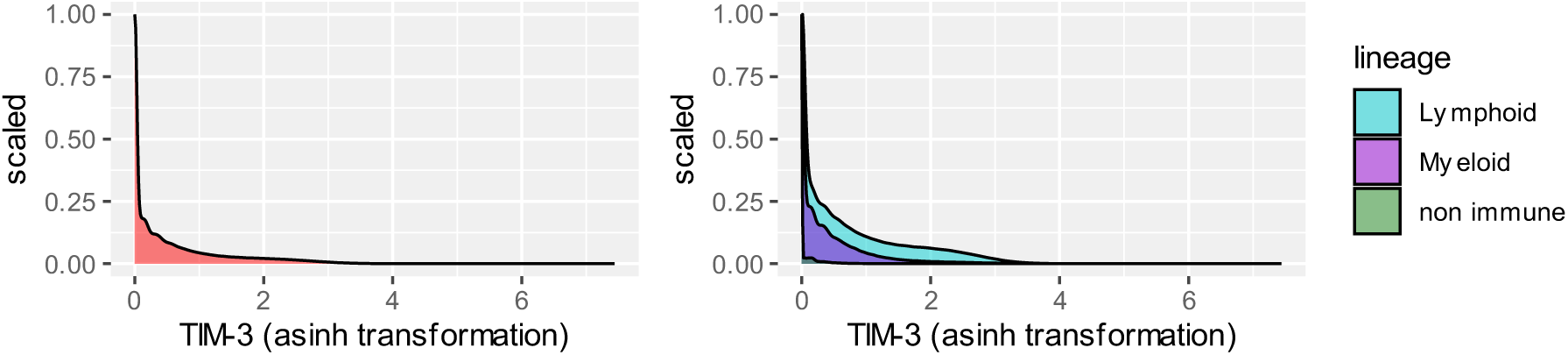
TIM-3 detection density plots on the mass cytometry dataset of Chevrier et al.. Values are asinh-transformed. Left: all cells. Right: By lineage

**Supp.Fig.17:**
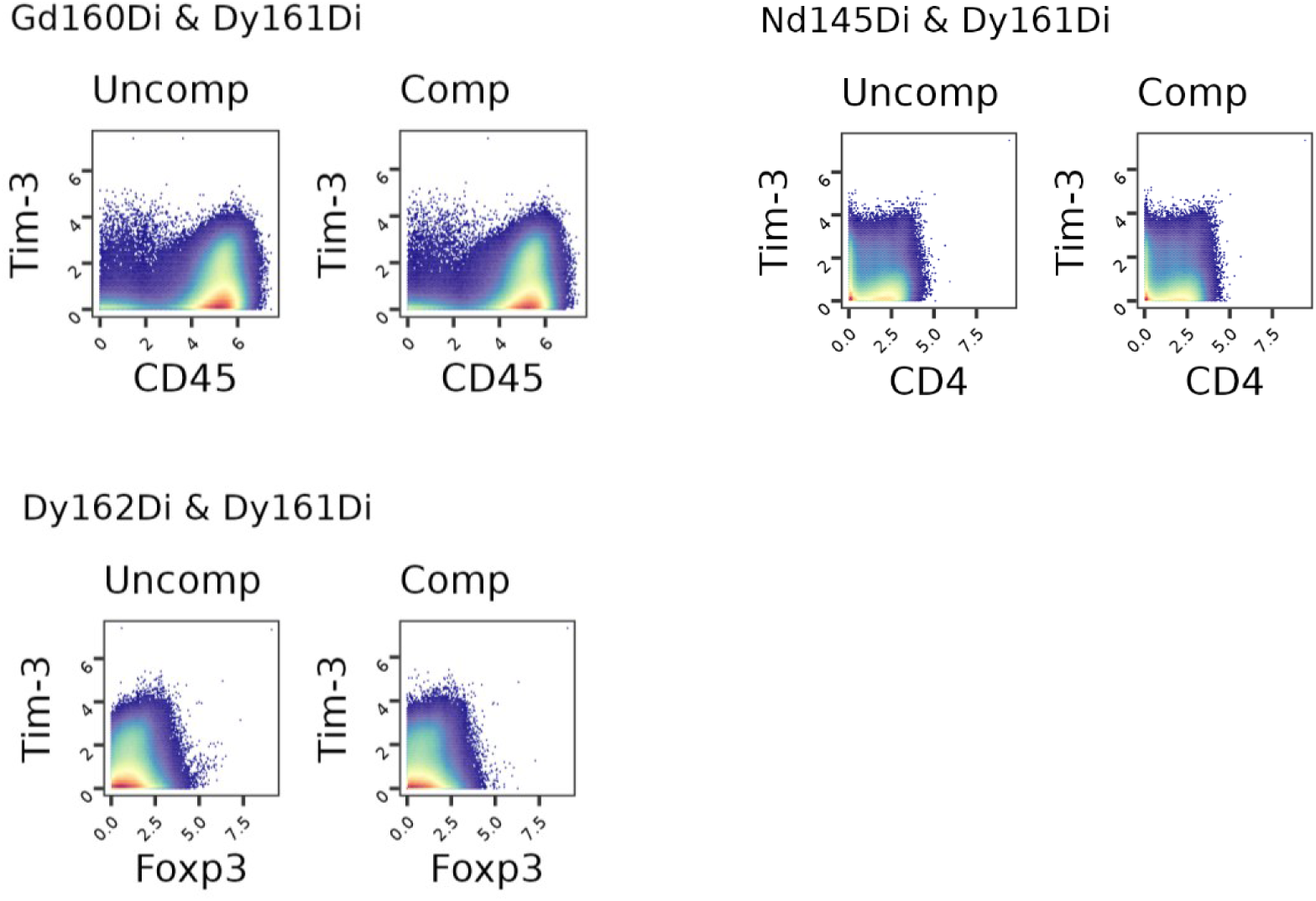
Contour plots visualizing the effect of spillover compensation on TIM-3 signal measure and potentially interacting mass channels. Values are asinh-transformed. “Uncomp” = uncompensated values. “Comp” = after compensation.

**Supp.Fig.18:**
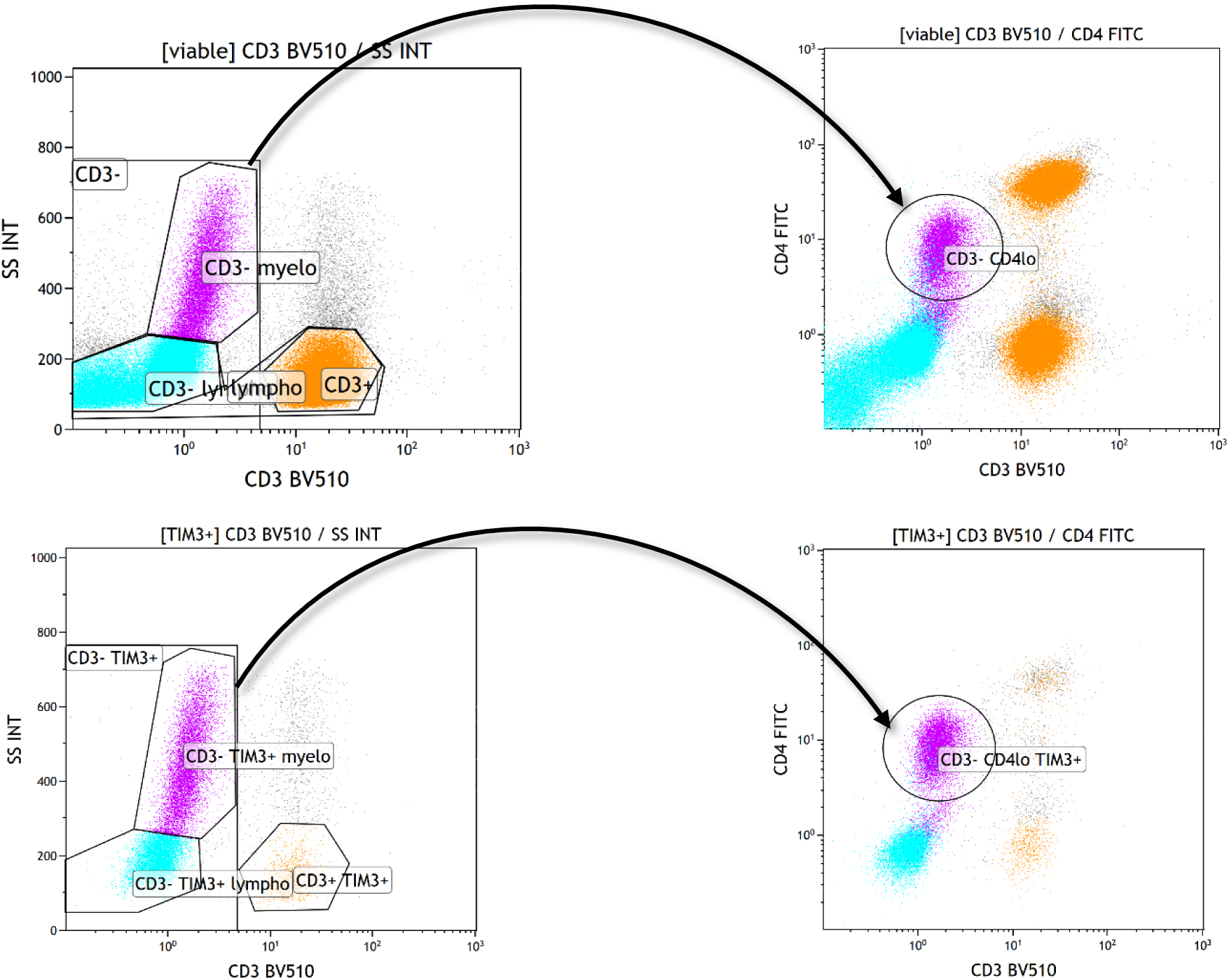
Representative flow cytometry plot showing intermediate levels of CD4 expression by the “CD3- myeloid” (top left and right, gated on viable cells) and “TIM3+CD3- myeloid” (bottom left and right, gated on TIM3+ cells) populations.

**Supp.Fig.19:**
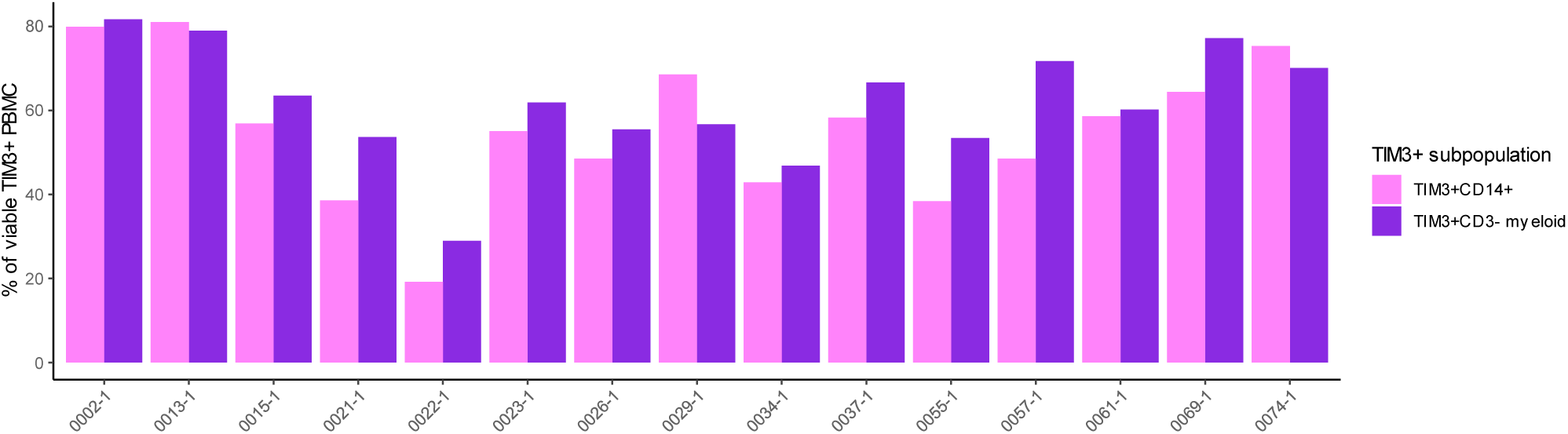
Head-to-head comparison of the CD14+ and CD3- myeloid cells proportions within TIM3+ PBMC, showing the correspondence of the two populations in a subset of patients from the Colcheckpoint cohort (n = 15), for which an additional cytometry panel with CD14 was performed on the same PBMC sample.

**Supp.Fig.20:**
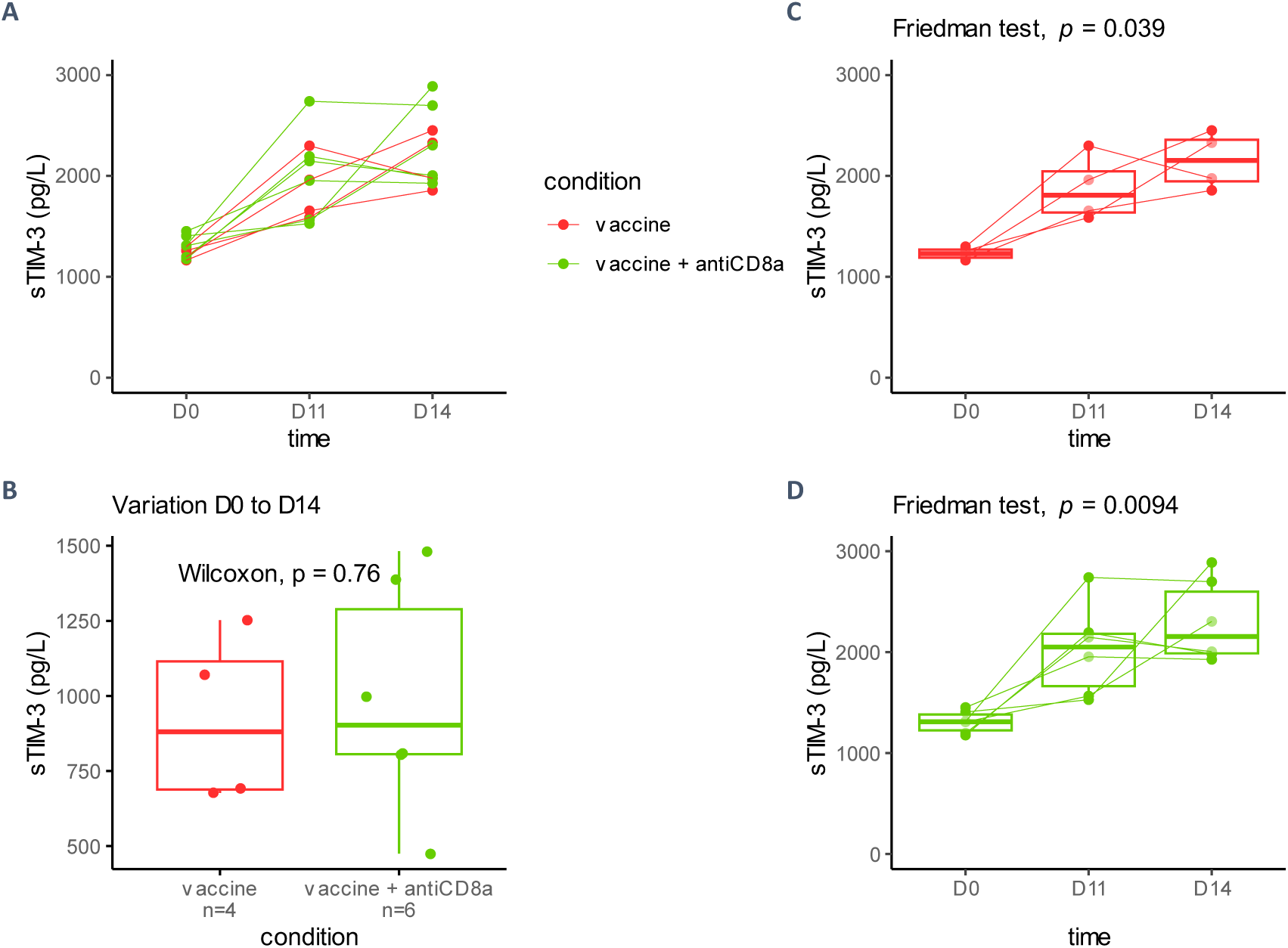
sTIM-3 plasmatic levels in syngeneic mice inoculated with TC-1 cells, immunized with an antitumor vaccine and ± depleted for CD8+ T cells. red: vaccinated (at D7, n= 4); green: vaccinated + anti-CD8α (vaccine at D7, anti-CD8α at D5 and D12; n= 5). Serum sTIM-3 measures are given in pg/L. **A.** All measures. **B.** Comparison of serum sTIM-3 variation between day 0 (D0, tumor graft) and day 14 (D14) in vaccinated versus vaccinated + CD8 T cells depleted mice. **C. & D**. serum sTIM-3 evolution over time in each group.

## Supplementary tables

**Supp.Table 1:**
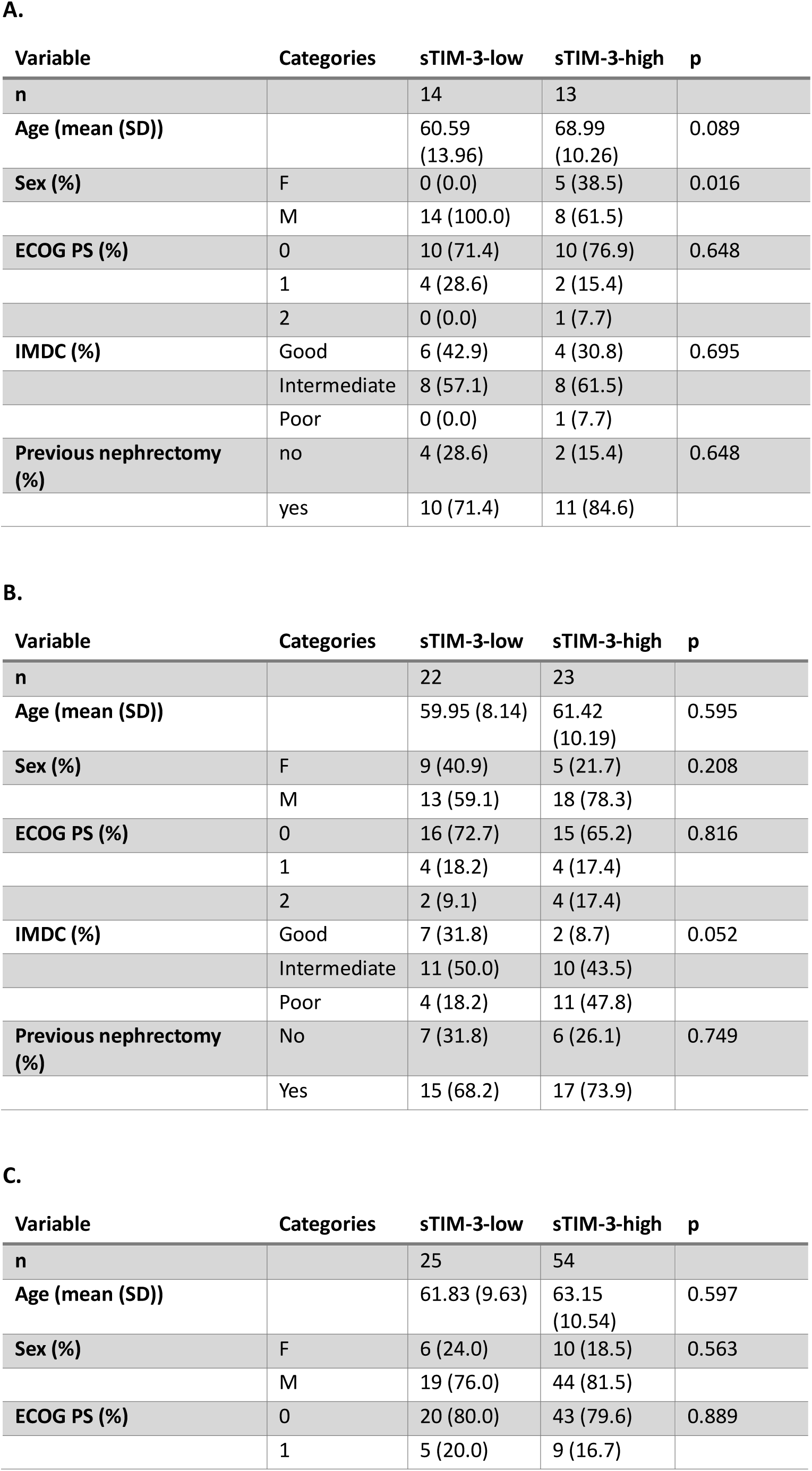
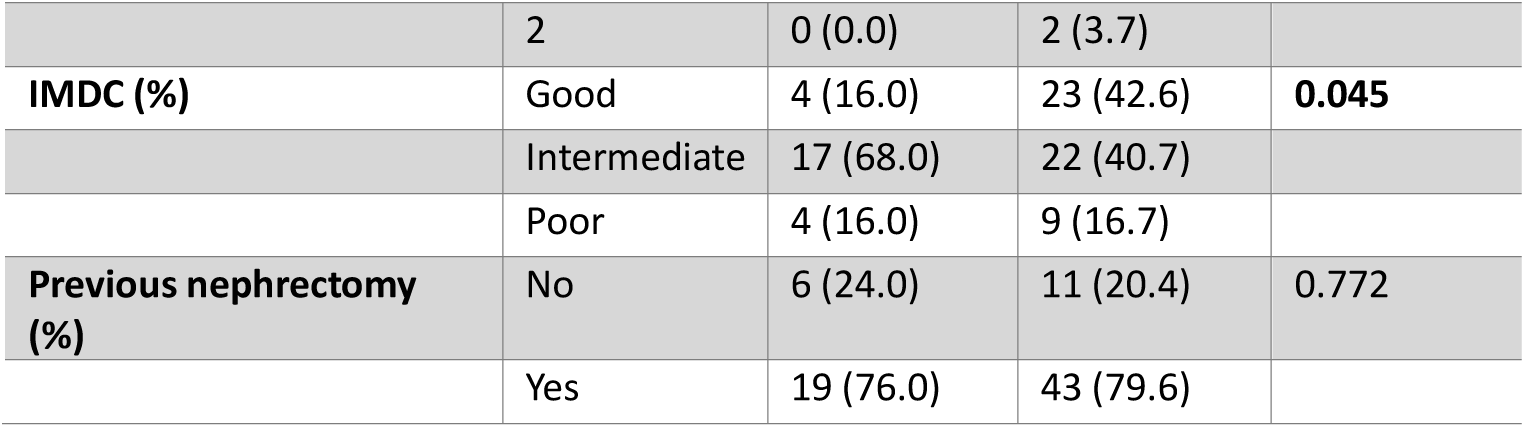
Characteristics of participants analyzed for OS, stratified on sTIM-3 categorization in **A.** Colcheckpoint. **B.** BIONIKK nivolumab-treated. **C.** BIONIKK nivolumab-ipilimumab treated.

| Variable | Categories | sTIM-3-low | sTIM-3-high | p |
| --- | --- | --- | --- | --- |
| **n** |  | 14 | 13 |  |
| **Age (mean (SD))** |  | 60.59<br>(13.96) | 68.99<br>(10.26) | 0.089 |
| **Sex (%)** | F | 0 (0.0) | 5 (38.5) | 0.016 |
|  | M | 14 (100.0) | 8 (61.5) |  |
| **ECOG PS (%)** | 0 | 10 (71.4) | 10 (76.9) | 0.648 |
|  | 1 | 4 (28.6) | 2 (15.4) |  |
|  | 2 | 0 (0.0) | 1 (7.7) |  |
| **IMDC (%)** | Good | 6 (42.9) | 4 (30.8) | 0.695 |
|  | Intermediate | 8 (57.1) | 8 (61.5) |  |
|  | Poor | 0 (0.0) | 1 (7.7) |  |
| **Previous nephrectomy (%)** | no | 4 (28.6) | 2 (15.4) | 0.648 |
|  | yes | 10 (71.4) | 11 (84.6) |  |

B.
| Variable | Categories | sTIM-3-low | sTIM-3-high | p |
| --- | --- | --- | --- | --- |
| **n** |  | 22 | 23 |  |
| **Age (mean (SD))** |  | 59.95 (8.14) | 61.42<br>(10.19) | 0.595 |
| **Sex (%)** | F | 9 (40.9) | 5 (21.7) | 0.208 |
|  | M | 13 (59.1) | 18 (78.3) |  |
| **ECOG PS (%)** | 0 | 16 (72.7) | 15 (65.2) | 0.816 |
|  | 1 | 4 (18.2) | 4 (17.4) |  |
|  | 2 | 2 (9.1) | 4 (17.4) |  |
| **IMDC (%)** | Good | 7 (31.8) | 2 (8.7) | 0.052 |
|  | Intermediate | 11 (50.0) | 10 (43.5) |  |
|  | Poor | 4 (18.2) | 11 (47.8) |  |
| **Previous nephrectomy (%)** | No | 7 (31.8) | 6 (26.1) | 0.749 |
|  | Yes | 15 (68.2) | 17 (73.9) |  |

C.
| Variable | Categories | sTIM-3-low | sTIM-3-high | p |
| --- | --- | --- | --- | --- |
| **n** |  | 25 | 54 |  |
| **Age (mean (SD))** |  | 61.83 (9.63) | 63.15<br>(10.54) | 0.597 |
| **Sex (%)** | F | 6 (24.0) | 10 (18.5) | 0.563 |
|  | M | 19 (76.0) | 44 (81.5) |  |
| **ECOG PS (%)** | 0 | 20 (80.0) | 43 (79.6) | 0.889 |
|  | 1 | 5 (20.0) | 9 (16.7) |  |
|  | 2 | 0 (0.0) | 2 (3.7) |  |
| **IMDC (%)** | Good | 4 (16.0) | 23 (42.6) | **0.045** |
|  | Intermediate | 17 (68.0) | 22 (40.7) |  |
|  | Poor | 4 (16.0) | 9 (16.7) |  |
| **Previous nephrectomy (%)** | No | 6 (24.0) | 11 (20.4) | **0.772** |
|  | Yes | 19 (76.0) | 43 (79.6) |  |

**Supp.Table 2:** Antibodies used for multiplex IHC and flow cytometry experiments. **A.** IHC antibodies. **B.** Flow cytometry antibodies. Staining was performed with 3 separate panels: 1^st^ panel: anti-CD3, anti- CD4, anti-TIM-3; 2^nd^ panel: anti-CD3, anti-CD8, anti-TIM-3; 3^rd^ panel: anti- CD14, anti-CD19, anti-TIM- 3.

**A.**
| Target | Manufacturer | Reference | Native concentration | Dilution |
| --- | --- | --- | --- | --- |
| **TIM-3** | CST® | 45208 | 16 µg/mL | 1:300 |
| **PAX8** | Abcam® | ab191870 | 853 µg/mL | 1:400 |
| **pan-Cytokeratin** | CST® | 4545 | 190 µg/mL | 1:250 |

**B.**
| Target | Manufacturer | Reference | Fluorophore |
| --- | --- | --- | --- |
| **CD3** | BioLegend® | 300448 | BV510 |
| **CD4** | BioLegend® | 300506 | FITC |
| **TIM-3** | BioLegend® | 345006 | PE |
| **CD8** | BioLegend® | 301006 | FITC |
| **CD14** | BioLegend® | 367124 | BV510 |
| **CD19** | BioLegend® | 302206 | FITC |

## Acknowledgment

The authors thank Marine Sroussi, Aurélien De Reynies, Pierre Gestraud, Lorette Noiret and Aleksandar Obradovic for their discussions and advice on scRNAseq data analyzes; Stéphane Chevrier for his advice on mass cytometry data exploitation; Florian Da Silva and Chloé Broudin for their help on tumor samples logistics; Jean-Philippe Empana for his advice on survival models; Clémence Granier for inspiring scientific discussions.

## Data sharing statement

Requests for anonymized patient data will be examined on an individual basis by relevant administrating committees of the Colcheckpoint and BIONIKK cohorts data. Processed data from animal experiments are available on request from the corresponding authors.

## References

1. FDA C for DE and. Approved Drugs - Nivolumab (Opdivo Injection) - advanced renal cell carcinoma [Internet]. Center for Drug Evaluation and Research 2015 [cited September 18, 2023]. Available at: http://wayback.archive-it.org/7993/20170111231614/http://www.fda.gov/Drugs/InformationOnDrugs/ApprovedDrugs/ucm474092.htm

2. Powles T, Albiges L, Bex A, Comperat E, Grünwald V, Kanesvaran R, et al. Renal cell carcinoma: ESMO Clinical Practice Guideline for diagnosis, treatment and follow-up. Ann Oncol. May 2024;S0923753424006768.

3. Heng DY, Xie W, Regan MM, Harshman LC, Bjarnason GA, Vaishampayan UN, et al. External validation and comparison with other models of the International Metastatic Renal-Cell Carcinoma Database Consortium prognostic model: a population-based study. Lancet Oncol. February 1, 2013;14(2):141–148.

4. Motzer RJ, Rini BI, McDermott DF, Arén Frontera O, Hammers HJ, Carducci MA, et al. Nivolumab plus ipilimumab versus sunitinib in first-line treatment for advanced renal cell carcinoma: extended follow-up of efficacy and safety results from a randomised, controlled, phase 3 trial. Lancet Oncol. October 2019;20(10):1370–1385.

5. Tang R, Rangachari M, Kuchroo VK. Tim-3: A co-receptor with diverse roles in T cell exhaustion and tolerance. Semin Immunol. April 1, 2019;42:101302.

6. Granier C, Dariane C, Combe P, Verkarre V, Urien S, Badoual C, et al. Tim-3 Expression on Tumor-Infiltrating PD-1+CD8+ T Cells Correlates with Poor Clinical Outcome in Renal Cell Carcinoma. Cancer Res. March 1, 2017;77(5):1075–1082.

7. Hu J, Chen Z, Bao L, Zhou L, Hou Y, Liu L, et al. Single-Cell Transcriptome Analysis Reveals Intratumoral Heterogeneity in ccRCC, which Results in Different Clinical Outcomes. Mol Ther. July 8, 2020;28(7):1658–1672.

8. Pignon J-C, Jegede O, Shukla SA, Braun DA, Horak CE, Wind-Rotolo M, et al. irRECIST for the Evaluation of Candidate Biomarkers of Response to Nivolumab in Metastatic Clear Cell Renal Cell Carcinoma: Analysis of a Phase II Prospective Clinical Trial. Clin Cancer Res Off J Am Assoc Cancer Res. 2019;25(7):2174–2184.

9. Ficial M, Jegede OA, Sant’Angelo M, Hou Y, Flaifel A, Pignon J-C, et al. Expression of T-Cell Exhaustion Molecules and Human Endogenous Retroviruses as Predictive Biomarkers for Response to Nivolumab in Metastatic Clear Cell Renal Cell Carcinoma. Clin Cancer Res. March 1, 2021;27(5):1371–1380.

10. Saliby RM, El Zarif T, Bakouny Z, Shah V, Xie W, Flippot R, et al. Circulating and Intratumoral Immune Determinants of Response to Atezolizumab plus Bevacizumab in Patients with Variant Histology or Sarcomatoid Renal Cell Carcinoma. Cancer Immunol Res. August 3, 2023;11(8):1114–1124.

11. Tian T, Li Z. Targeting Tim-3 in Cancer With Resistance to PD-1/PD-L1 Blockade. Front Oncol. 2021;11:731175.

12. Cai L, Li Y, Tan J, Xu L, Li Y. Targeting LAG-3, TIM-3, and TIGIT for cancer immunotherapy. J Hematol OncolJ Hematol Oncol. September 5, 2023;16(1):101.

13. Möller-Hackbarth K, Dewitz C, Schweigert O, Trad A, Garbers C, Rose-John S, et al. A disintegrin and metalloprotease (ADAM) 10 and ADAM17 are major sheddases of T cell immunoglobulin and mucin domain 3 (Tim-3). J Biol Chem. November 29, 2013;288(48):34529–34544.

14. Clayton KL, Douglas-Vail MB, Nur-ur Rahman AKM, Medcalf KE, Xie IY, Chew GM, et al. Soluble T cell immunoglobulin mucin domain 3 is shed from CD8+ T cells by the sheddase ADAM10, is increased in plasma during untreated HIV infection, and correlates with HIV disease progression. J Virol. April 2015;89(7):3723–3736.

15. Giraldo NA, Becht E, Vano Y, Petitprez F, Lacroix L, Validire P, et al. Tumor-Infiltrating and Peripheral Blood T-cell Immunophenotypes Predict Early Relapse in Localized Clear Cell Renal Cell Carcinoma. Clin Cancer Res Off J Am Assoc Cancer Res. August 1, 2017;23(15):4416–4428.

16. Wang Q, Zhang J, Tu H, Liang D, Chang DavidW, Ye Y, et al. Soluble immune checkpoint-related proteins as predictors of tumor recurrence, survival, and T cell phenotypes in clear cell renal cell carcinoma patients. J Immunother Cancer. November 29, 2019;7(1):334.

17. Vano Y-A, Elaidi R, Bennamoun M, Chevreau C, Borchiellini D, Pannier D, et al. Nivolumab, nivolumab–ipilimumab, and VEGFR-tyrosine kinase inhibitors as first-line treatment for metastatic clear-cell renal cell carcinoma (BIONIKK): a biomarker-driven, open-label, non-comparative, randomised, phase 2 trial. Lancet Oncol. May 2022;23(5):612–624.

18. Obradovic A, Chowdhury N, Haake SM, Ager C, Wang V, Vlahos L, et al. Single-cell protein activity analysis identifies recurrence-associated renal tumor macrophages. Cell. May 2021;184(11):2988–3005.e16.

19. Bi K, He MX, Bakouny Z, Kanodia A, Napolitano S, Wu J, et al. Tumor and immune reprogramming during immunotherapy in advanced renal cell carcinoma. Cancer Cell. May 10, 2021;39(5):649–661.e5.

20. Borcherding N, Vishwakarma A, Voigt AP, Bellizzi A, Kaplan J, Nepple K, et al. Mapping the immune environment in clear cell renal carcinoma by single-cell genomics. Commun Biol. January 27, 2021 [cited February 15, 2021];4. Available at: https://www.ncbi.nlm.nih.gov/pmc/articles/PMC7840906/

21. Braun DA, Street K, Burke KP, Cookmeyer DL, Denize T, Pedersen CB, et al. Progressive immune dysfunction with advancing disease stage in renal cell carcinoma. Cancer Cell. May 2021;39(5):632–648.e8.

22. Stewart BJ, Ferdinand JR, Young MD, Mitchell TJ, Loudon KW, Riding AM, et al. Spatio-temporal immune zonation of the human kidney. Science. September 27, 2019;365(6460):1461–1466.

23. Liu X, Han W, Hu X. Chapter Three - Post-transcriptional regulation of myeloid cell-mediated inflammatory responses. In Alt FW, Murphy KM (eds): Advances in Immunology Academic Press 2023 [cited December 18, 2023]; 59–82. (Advances in Immunology; vol. 160). Available at: https://www.sciencedirect.com/science/article/pii/S0065277623000299

24. Komohara Y, Morita T, Annan DA, Horlad H, Ohnishi K, Yamada S, et al. The Coordinated Actions of TIM-3 on Cancer and Myeloid Cells in the Regulation of Tumorigenicity and Clinical Prognosis in Clear Cell Renal Cell Carcinomas. Cancer Immunol Res. September 1, 2015;3(9):999–1007.

25. Gonçalves Silva I, Yasinska IM, Sakhnevych SS, Fiedler W, Wellbrock J, Bardelli M, et al. The Tim- 3-galectin-9 Secretory Pathway is Involved in the Immune Escape of Human Acute Myeloid Leukemia Cells. eBioMedicine. August 1, 2017;22:44–57.

26. Nielsen R, Christensen EI, Birn H. Megalin and cubilin in proximal tubule protein reabsorption: from experimental models to human disease. Kidney Int. January 1, 2016;89(1):58–67.

27. Sabatos CA, Chakravarti S, Cha E, Schubart A, Sánchez-Fueyo A, Zheng XX, et al. Interaction of Tim-3 and Tim-3 ligand regulates T helper type 1 responses and induction of peripheral tolerance. Nat Immunol. November 2003;4(11):1102–1110.

28. Dewitz C, Möller-Hackbarth K, Schweigert O, Reiss K, Chalaris A, Scheller J, et al. T-cell immunoglobulin and mucin domain 2 (TIM-2) is a target of ADAM10-mediated ectodomain shedding. FEBS J. 2014;281(1):157–174.

29. Fyfe G, Fisher RI, Rosenberg SA, Sznol M, Parkinson DR, Louie AC. Results of treatment of 255 patients with metastatic renal cell carcinoma who received high-dose recombinant interleukin-2 therapy. J Clin Oncol. March 1995;13(3):688–696.

30. Black RA, Rauch CT, Kozlosky CJ, Peschon JJ, Slack JL, Wolfson MF, et al. A metalloproteinase disintegrin that releases tumour-necrosis factor-α from cells. Nature. February 1997;385(6618):729–733.

31. Ivetic A, Hoskins Green HL, Hart SJ. L-selectin: A Major Regulator of Leukocyte Adhesion, Migration and Signaling. Front Immunol. May 14, 2019;10:1068.

32. Khantakova D, Brioschi S, Molgora M. Exploring the Impact of TREM2 in Tumor-Associated Macrophages. Vaccines. June 14, 2022;10(6):943.

33. Vanmeerbeek I, Naulaerts S, Sprooten J, Laureano RS, Govaerts J, Trotta R, et al. Targeting conserved TIM3+VISTA+ tumor-associated macrophages overcomes resistance to cancer immunotherapy. Sci Adv. July 19, 2024;10(29):eadm8660.

34. Magen A, Hamon P, Fiaschi N, Soong BY, Park MD, Mattiuz R, et al. Intratumoral dendritic cell–CD4+ T helper cell niches enable CD8+ T cell differentiation following PD-1 blockade in hepatocellular carcinoma. Nat Med. June 2023;29(6):1389–1399.

35. Chen JH, Nieman LT, Spurrell M, Jorgji V, Elmelech L, Richieri P, et al. Human lung cancer harbors spatially organized stem-immunity hubs associated with response to immunotherapy. Nat Immunol. April 2024;25(4):644–658.

36. Espinosa-Carrasco G, Chiu E, Scrivo A, Zumbo P, Dave A, Betel D, et al. Intratumoral immune triads are required for immunotherapy-mediated elimination of solid tumors. Cancer Cell. June 20, 2024 [cited July 2, 2024]; Available at: https://www.sciencedirect.com/science/article/pii/S1535610824001934

37. Roquilly A, Jacqueline C, Davieau M, Mollé A, Sadek A, Fourgeux C, et al. Alveolar macrophages are epigenetically altered after inflammation, leading to long-term lung immunoparalysis. Nat Immunol. June 2020;21(6):636–648.

38. Chevrier S, Levine JH, Zanotelli VRT, Silina K, Schulz D, Bacac M, et al. An Immune Atlas of Clear Cell Renal Cell Carcinoma. Cell. May 4, 2017;169(4):736–749.e18.

39. Chevrier S, Crowell HL, Zanotelli VRT, Engler S, Robinson MD, Bodenmiller B. Compensation of Signal Spillover in Suspension and Imaging Mass Cytometry. Cell Syst. May 23, 2018;6(5):612–620.e5.

